# Wastewater intelligence predicts the emergence of clinically-relevant and drug-resistant *Candida auris* at healthcare facilities

**DOI:** 10.1101/2025.07.29.25332388

**Authors:** Ching-Lan Chang, Michael A. Moshi, Quang-Huy Nguyen, Junghun Oh, Ha Nguyen, Pratik Paranjape, Mohammed Abushanab, Austin J. Tang, Jose Yani Itorralba, Lauryn Massic, Eakalak Khan, Cassius Lockett, Horng-Yuan Kan, Mark Pandori, Daniel Gerrity, Van Vo, Tin Nguyen, David Hess, Edwin C. Oh

**Affiliations:** Laboratory of Neurogenetics and Precision Medicine, College of Sciences; Neuroscience Interdisciplinary Ph.D. program; Department of Computer Science and Software Engineering, Auburn University, Auburn, AL 36849; Nevada State Public Health Laboratory, 1660 N Virginia St, Reno, NV 89503; Civil and Environmental Engineering and Construction Department; Southern Nevada Health District, Las Vegas NV, 89106; Southern Nevada Water Authority, P.O. Box 99954, Las Vegas NV, 89193, USA; Department of Brain Health; Department of Internal Medicine, Kirk Kerkorian School of Medicine at UNLV, University of Nevada Las Vegas, Las Vegas, NV 89154

**Keywords:** MALDI, amplicon sequencing, whole genome sequencing, multidrug-resistance, variants, wastewater-based epidemiology (WBE), antimicrobial resistance (AMR)

## Abstract

The rapid evolution of antifungal resistance in *Candida auris* presents significant challenges for conventional public health surveillance methods, particularly in detecting emergent and highly transmissible drug-resistant variants. Using wastewater-based epidemiology (WBE) tools initially developed during the COVID-19 pandemic, we implemented a high-resolution facility-level early warning system to monitor *C. auris* infections and resistance patterns. Our comprehensive evaluation across Southern Nevada demonstrated that upstream sewage monitoring at healthcare facilities provided significant sensitivity (p<0.001) compared to wastewater treatment plant (WWTP) sampling. By combining amplicon sequencing and MALDI-TOF mass spectrometry, we identified clinically relevant resistance-associated variants in wastewater samples, while whole genome sequencing revealed >90% genomic concordance between 443 wastewater-derived genomes and 2,977 clinical isolates. We also detected novel subclades and resistance mutations, including *FKS1* Phe635Leu and co-occurring *ERG11*/*FKS1* variants in wastewater samples up to nearly five months before their appearance in clinical settings. Further transcriptomic profiling of drug-resistant isolates under antifungal and stress conditions identified previously uncharacterized adaptation mechanisms, including differential regulation of ribosomal assembly pathways and cell cycle checkpoints. These findings highlight how wastewater intelligence can substantially enhance traditional public health surveillance approaches for the early and proactive detection and monitoring of *C. auris* outbreaks and antifungal resistance.

## Introduction

The global emergence of *Candida auris* in healthcare settings represents an unprecedented and complex combination of rapid antifungal resistance development and remarkable environmental adaptability^1–7^. Since its identification in 2009, *C. auris* has rapidly evolved resistance to multiple antifungal drug classes, including azoles (e.g., fluconazole), echinocandins (e.g., micafungin), and polyenes (e.g., amphotericin B), leading to its designation as the only fungal organism classified as an urgent public health threat by the United States (U.S.) Centers for Disease Control and Prevention (CDC)^8^. In healthcare settings, this pathogen causes severe infections with mortality rates of up to 30-60% among infected patients^9^. The infections show a distinct demographic pattern, predominantly affecting older adults, especially those with compromised immune systems or those requiring indwelling medical devices, such as catheters or breathing tubes^10,11^. The pathogen’s ability to develop resistance through multiple mechanisms, particularly mutations in ergosterol biosynthesis and drug efflux pathways, has severely constrained treatment options and highlighted the need for development of novel surveillance strategies^12,13^.

The global dissemination of *C. auris* is characterized by six distinct geographical clades exhibiting unique resistance profiles and evolutionary trajectories^4,5,14^. Clade I isolates, which originated in South Asia, demonstrate highly prevalent rates of azole resistance (particularly to fluconazole) and enhanced transmissibility within healthcare settings. Clade II, first identified in East Asia, typically shows lower resistance rates to all three major antifungal classes while maintaining fitness in healthcare environments. Clade III strains, initially detected in Africa, exhibit heterogeneous susceptibility patterns, including reduced sensitivity to amphotericin B and moderate fluconazole resistance. Clade IV, emerging from South America, shows moderate levels of antifungal resistance, particularly to fluconazole and occasionally to echinocandins. Clade V, identified in Iran, represents a recently characterized lineage with high resistance to multiple antifungal classes. The newest addition, Clade VI, was first detected in Singapore and Bangladesh and shares the closest genetic relationships with Clade IV, but remains distinctly separated by at least 37,000 SNPs and demonstrates variable antifungal susceptibility patterns. These clade-specific variations in resistance mechanisms and virulence factors underscore the complexity of tracking and responding to emerging antifungal resistance. Understanding these distinct evolutionary patterns has become increasingly relevant for developing precision-driven surveillance and treatment strategies, particularly as new variants continue to emerge.

Recent advances in high-resolution genomic epidemiology have revealed key molecular mechanisms underlying *C. auris* resistance, including clinically significant mutations in *ERG11* (Tyr132Phe) and *FKS1* (Ser639Phe) that confer resistance to azoles and echinocandins, respectively^13^. However, traditional clinical surveillance methods face inherent challenges in rapidly detecting and responding to new outbreaks and emerging resistance patterns. These limitations stem from the baseline timeframe for an infection to present clinically and undergo confirmatory testing, compounded by reliance on timely and active participation from healthcare facilities, which may be reluctant to engage in expanded surveillance. Such delays or outright refusals to participate create critical opportunities for resistant strains to disseminate within healthcare networks^15^. Recent efforts to leverage wastewater-based epidemiology (WBE) have demonstrated the feasibility of detecting *C. auris* DNA in wastewater treatment plants (WWTPs) across the U.S., establishing a promising foundation for community-level surveillance^16–20^. Despite these new approaches to surveillance of bacterial, fungal, and viral pathogens^21–23^, there is still a critical need to link wastewater findings to facility-specific and/or community-level transmission dynamics to further increase the actionability of wastewater intelligence data.

Here, we demonstrate that integrated wastewater intelligence provides early and actionable warning of emerging *C. auris* subclades during the 2022-2024 outbreak in Las Vegas, Nevada—the largest recorded outbreak in U.S. history to date^24,25^. Through comprehensive genomic, transcriptomic, and phenotypic analyses, we establish that wastewater monitoring, particularly at healthcare facilities, enables detection of emerging subclades up to nearly five months before their appearance in clinical isolates. Our comparative analysis reveals remarkable 90% genomic concordance between wastewater and clinical isolates, while uncovering novel stress response mechanisms and metabolic adaptations associated with antifungal resistance. These findings demonstrate the power of wastewater surveillance in tracking fungal pathogen evolution and resistance patterns, providing a robust framework for proactive infection monitoring and potentially guiding treatment strategies.

## Results

### Epidemiological and molecular surveillance reveals distinct patterns of *C. auris* transmission

Longitudinal clinical surveillance of hospitals and long-term care facilities in Southern Nevada from August 2021 to August 2024 revealed a significant outbreak pattern, with a marked increase in confirmed colonizations and clinical cases beginning in the spring of 2022 (**Figure 1A**). During this time, reported cases surged above the baseline rates observed in 2021/early 2022, reaching cumulative totals of 1,461 clinical cases and 2,738 colonizations by August 2024^26^. Demographic analysis showed distinct patterns in the affected population up to February 2024, with a slight male predominance (58% male versus 40% female, 2% not reported)^27^ (**Figure 1B**). Age stratification demonstrated the outbreak’s disproportionate impact on older adults, with the majority of cases occurring in individuals over 50 years of age. Peak incidence was observed in those over 70 years (45.4%), followed by the 60-69 age group (28.7%)^27^, which is consistent with the previously documented vulnerability of older populations to *C. auris* infection^10^.

**Figure 1.**
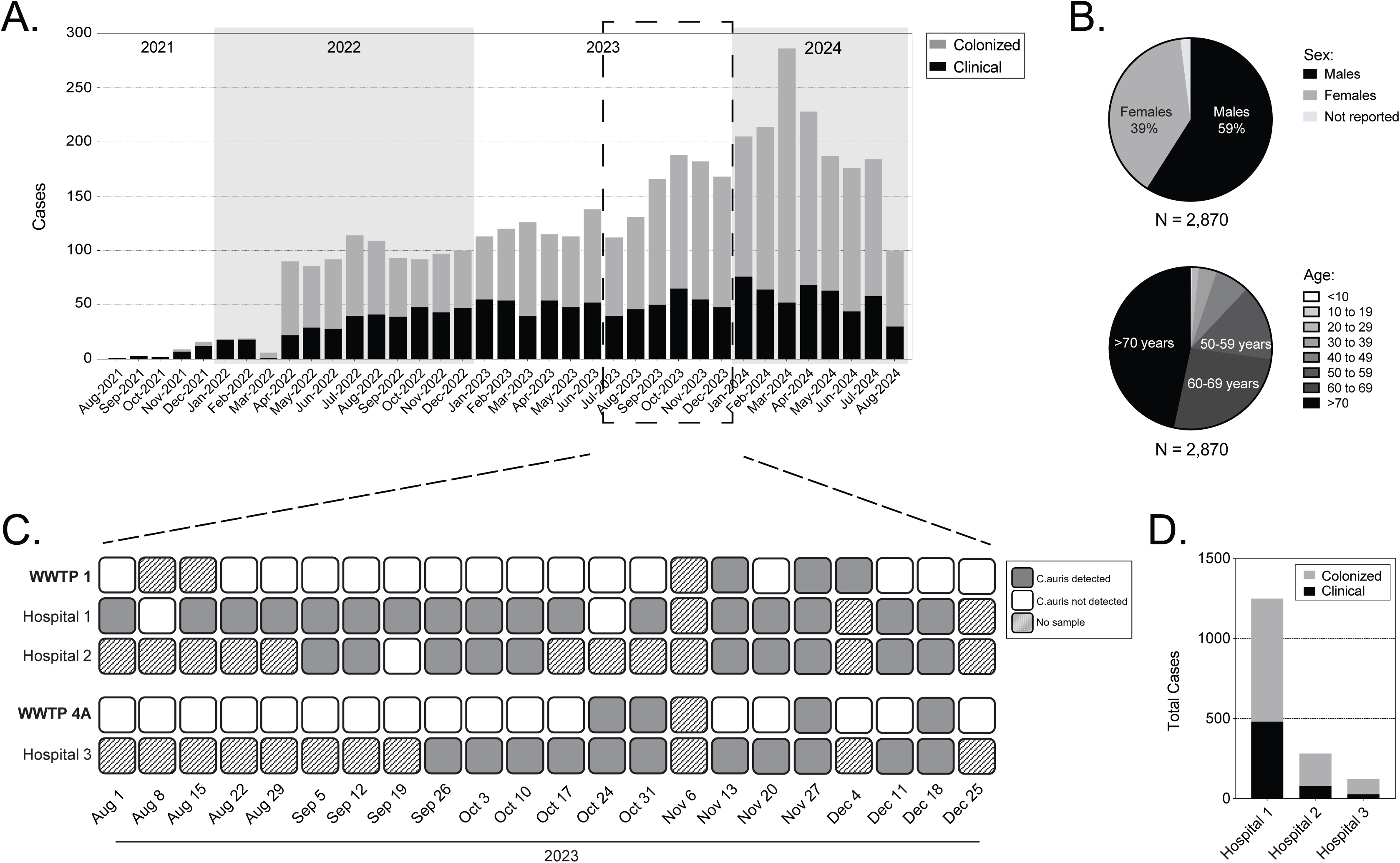
Characterization of *C. auris* outbreak dynamics through integrated clinical and wastewater surveillance approaches in Southern Nevada. **(A)** Monthly distribution of confirmed clinical cases (black) and colonization cases (gray) showing outbreak progression from August 2021 to August 2024, with a notable peak in late 2023/early 2024. **(B)** Pie chart demographic analysis of cases showing **(top)** sex distribution (2% not reported) and **(bottom)** age distribution (majority > 60 years). **(C)** Wastewater surveillance results from three hospitals and two wastewater treatment plants (WWTP 1 and 4A) between August-December 2023. Solid gray squares indicate *C. auris* detection, white squares indicate no detection, and hatched squares indicate no sample was collected on that date. **(D)** Total confirmed colonizations (gray) and clinical cases (black) at three major hospitals from 2021-2024.

To investigate whether wastewater surveillance could serve as a sensitive indicator of pathogen transmission within individual facilities, we conducted systematic wastewater monitoring at three major hospitals and their corresponding municipal WWTPs (WWTP1 and WWTP4A) between August and December 2023 (**Figure 1C**). This parallel sampling approach demonstrated that *C. auris* DNA concentrations in hospital wastewater were significantly correlated with the clinical case and colonization burden at each healthcare facility (p < 0.001). While *C. auris* DNA was successfully detected at both WWTPs (**Figure 1C**), the hospital wastewater signal was significantly more consistent (detection frequency = 92% vs. 17%) and the concentrations were significantly higher (p < 0.001, **Supplementary Figures 1-2**). In fact, the maximum concentrations in the hospital wastewater were nearly two orders of magnitude higher than the maximum concentrations at the community-scale WWTPs (∼10^8^ gc/L vs. ∼10^6^ gc/L). This dilution effect highlights the enhanced sensitivity and practical utility of facility-level surveillance for early outbreak detection. Analysis of case distribution across the three hospitals revealed distinct spatial transmission patterns, with Hospital 1 experiencing the highest cumulative burden (n = 1,249 cases), followed by Hospital 2 (n = 281) and Hospital 3 (n = 122)^26^ (**Figure 1D**).

### Targeted amplicon sequencing reveals dynamic *C. auris* populations rather than persistent wastewater biofilms

A fundamental question raised by prior *C. auris* wastewater surveillance studies is whether detection of this biofilm-associated pathogen represents active transmission or potentially just a persistent biofilm within the premise plumbing and/or sewer system^16^. To distinguish between these possibilities, we designed and implemented an amplicon panel targeting 11 genes in *C. auris* implicated in antifungal resistance mechanisms: *CDR1*, *ERG3*, *ERG6*, *ERG11*, *FKS1*, *HSP90*, *MEC3*, *MLH1*, *MRR1A*, *TAC1B*, and *UPC2* (**Supplementary Table 1**). To check for species discrimination in complex wastewater matrices, we identified and incorporated unique sequence markers for both *C. auris* and *C. albicans*. This targeted approach enabled spatial and temporal monitoring of resistance-associated variants across the three hospitals (**Figure 2**).

**Figure 2.**
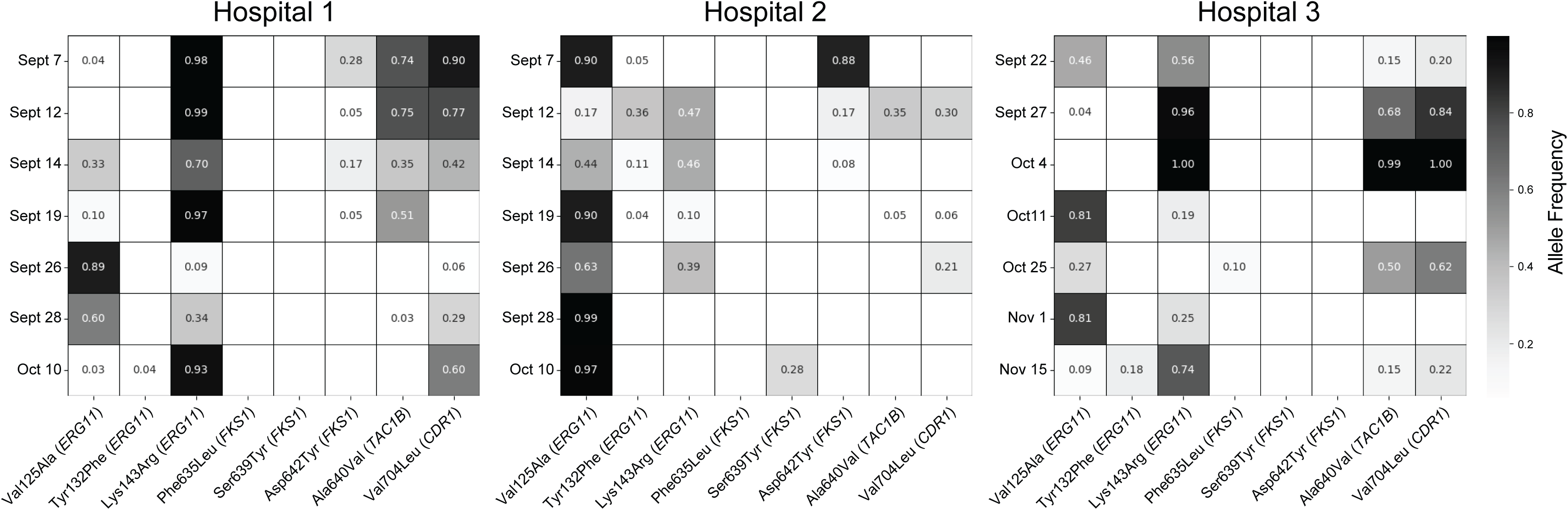
Spatial and temporal monitoring of resistance-associated variant frequencies at three hospitals between September 2023 and November 2023. Heatmaps show allele frequencies of mutations in genes associated with antifungal resistance: *ERG11* (azole resistance), *FKS1* (echinocandin resistance), *CDR1* and *TAC1B* (multidrug efflux). Colors represent variant allele frequencies from non-detect (white) to >75% (black). Hospital 1 (left panel) and Hospital 2 (middle panel) show emergence of *FKS1* Asp642Tyr independently. Hospital 2 (middle panel) displays rapid shifts in *ERG11* variants (Tyr132Phe and Lys143Arg) between September 7^th^ and September 19^st^ of 2023, and dynamic changes in multiple resistance markers over the sampling period. The temporal variation in mutation frequencies across all three facilities provides evidence for active transmission rather than stable biofilm colonization of wastewater systems.

High-resolution temporal analysis revealed rapid and independent fluctuations in variant frequencies that provided compelling evidence for active pathogen transmission rather than static biofilm persistence. For example, in Hospital 2, we observed dramatic shifts in *ERG11* variants Val125Ala and Lys143Arg, with allele frequencies surging from <10% to >75% between September 7 and September 19, 2023 (**Figure 2**). These abrupt changes occurred too rapidly to be explained by biofilm evolution alone. Supporting this interpretation, we detected the emergence of the *FKS1* Asp642Tyr mutation (associated with echinocandin resistance) in Hospitals 1 and 2 at different frequencies and timepoints. While patient movement between facilities could not be definitively tracked, the distinct temporal patterns and variant frequencies at each hospital suggest independent evolution under similar selective pressures rather than the spread of a single resistant clone.

Validation of our detection approach included parallel monitoring of species-specific sequences from both *C. auris* and *C. albicans* (**Supplementary Table 1**). The dual-panel design achieved reliable species discrimination with a detection sensitivity of 3% variant frequency in complex wastewater matrices, enabling identification of minor variants before they achieved dominance. We consistently detected multiple resistance-associated mutations, including the clinically significant *ERG11* Val125Ala and *CDR1* Val704Leu variants, demonstrating robust performance in wastewater samples. In addition, the *CDR1*, *ERG11*, *FKS1*, and *TAC1B* variants exhibited distinct facility-specific patterns of emergence and progression. These spatially resolved evolutionary trajectories showed unique patterns at each hospital, with variant frequencies changing independently over time. The facility-specific nature of these changes, combined with their rapid temporal dynamics, provided strong evidence that wastewater detection captured active *C. auris* transmission rather than persistent biofilm populations. This result is further supported by subsequent genomic analysis showing concordance between these wastewater-detected mutations and clinical isolates from the same facilities (discussed in detail below).

### MALDI-TOF mass spectrometry enables *C. auris* identification in complex wastewater matrices

To address the pressing need for accurate pathogen detection in healthcare settings, we developed and validated a MALDI-TOF mass spectrometry workflow for identifying *C. auris* in wastewater samples. We established comprehensive reference spectral libraries using characterized Clade I and Clade III isolates (**Figure 3A-B**). These reference spectra revealed distinctive diagnostic peaks across the 2,000-9,000 m/z range, with robust signals at approximately 6,000 m/z for both clades. Although sharing core spectral features, the clades exhibited unique peak patterns in the 6,000-7,000 m/z region, enabling clade differentiation. Control wastewater samples (confirmed negative for *C. auris* by qPCR) showed consistent background peaks at 3,000 m/z, characteristic of the organic matrix (**Figure 3C**).

**Figure 3.**
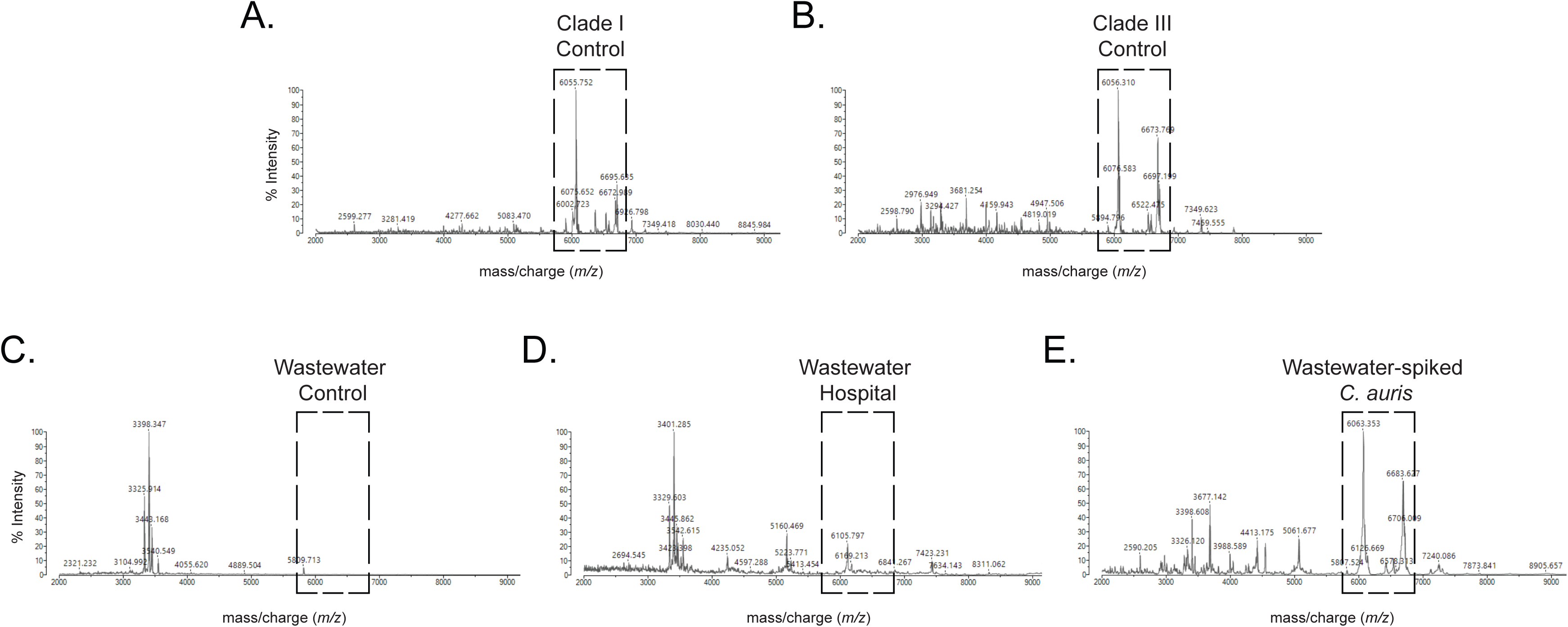
MALDI-TOF mass spectrometry for accurate *C. auris* detection in complex wastewater matrices. **(A)** Reference mass spectra from purified Clade I *C. auris* control isolate showing characteristic peaks in the 2,000-9,000 m/z range, with diagnostic signals at 6,000-7,000 m/z. **(B)** Reference mass spectra from Clade III *C. auris* control isolate displaying shared core spectral features with Clade I but exhibiting distinct peak patterns in the 6,000-7,000 m/z region, enabling clade discrimination. **(C)** Background mass spectra from control (non-spiked) wastewater showing low-intensity peaks around 3,000 m/z, establishing the baseline signature of the wastewater matrix. **(D)** Direct analysis of non-spiked hospital wastewater samples showing preservation of *C. auris*-specific spectral patterns, particularly in the 6,000-7,000 m/z range, validating direct detection of indigenous *C. auris* without pre-enrichment steps. **(E)** Mass spectra from wastewater samples spiked with known quantities of *C. auris*, demonstrating clear detection of pathogen-specific peaks (signal-to-noise ratio 2:1) against the background matrix.

Following a 24-48 h pre-enrichment step, hospital wastewater samples displayed *C. auris*-specific spectral patterns matching our reference isolates, with particularly strong signal preservation in the diagnostically crucial 6,000-7,000 m/z range, alongside the expected background signals at 3,000 m/z (**Figure 3D**). Systematic sensitivity analyses with wastewater samples spiked with defined quantities (∼10^6^ gc/L) of *C. auris* colonies (**Figure 3E**) achieved a signal-to-noise ratio of 2:1 for diagnostic peaks, confirming robust detection of *C. auris* in complex wastewater matrices. The remarkable stability of these high molecular weight peaks demonstrated minimal matrix interference in critical mass ranges. This optimized MALDI-TOF protocol provides definitive identification of *C. auris* in environmental samples, complementing existing molecular methods. Our findings establish MALDI-TOF mass spectrometry as a powerful tool for real-time pathogen surveillance in healthcare and wastewater/environmental settings, enabling rapid implementation of infection control measures.

### High genomic concordance between wastewater and clinical *C. auris* isolates enables early detection of emerging subclades

To evaluate whether wastewater surveillance could provide advance warning of emerging resistant variants, we conducted comprehensive genomic surveillance across Southern Nevada from August 2021 to May 2024. This systematic molecular approach characterized 2,977 clinical isolates from 45 healthcare facilities and 443 wastewater isolates from hospital wastewater and WWTPs. Species-level validation confirmed 98.9% (2,945/2,977) of clinical and 100.0% (443/443) of wastewater isolates as *C. auris*. Initial phylogenetic analysis revealed structured populations dominated by Clade I (31.4%, 934/2,977) and Clade III (67.7%, 2,014/2,977), with a single Clade IV isolate detected, suggesting multiple independent introductions in this geographic region (**Table 1**).

**Table 1.**
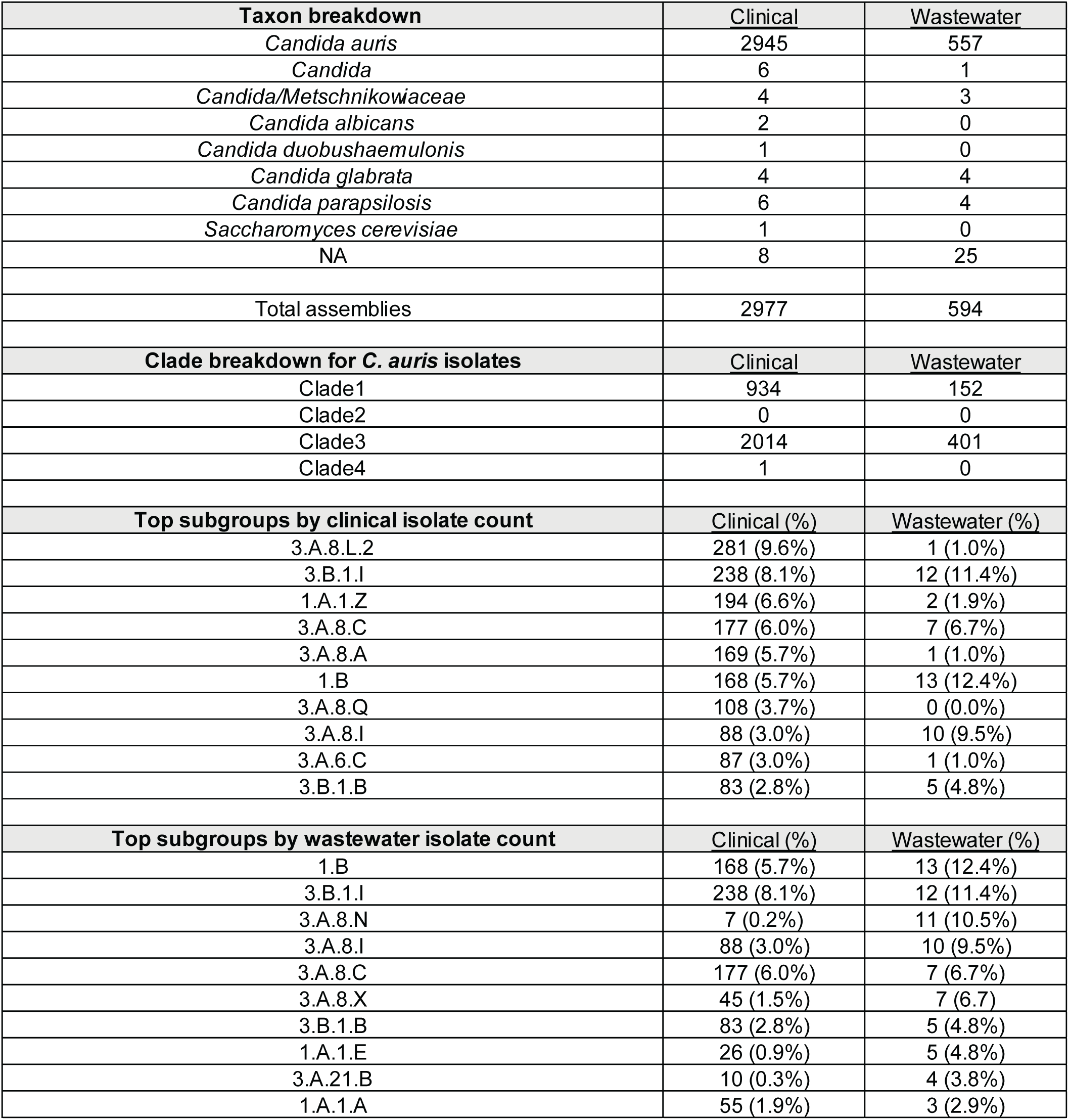
General description of clinical and wastewater isolates.

| **Taxon breakdown** | **Clinical** | **Wastewater** |
| --- | --- | --- |
| *Candida auris* | 2945 | 557 |
| *Candida* | 6 | 1 |
| *Candida/Metschnikowiaceae* | 4 | 3 |
| *Candida albicans* | 2 | 0 |
| *Candida duobushaemulonis* | 1 | 0 |
| *Candida glabrata* | 4 | 4 |
| *Candida parapsilosis* | 6 | 4 |
| *Saccharomyces cerevisiae* | 1 | 0 |
| NA | 8 | 25 |
| Total assemblies | 2977 | 594 |
| **Clade breakdown for *C. auris* isolates** | **Clinical** | **Wastewater** |
| Clade1 | 934 | 152 |
| Clade2 | 0 | 0 |
| Clade3 | 2014 | 401 |
| Clade4 | 1 | 0 |
| **Top subgroups by clinical isolate count** | **Clinical (%)** | **Wastewater (%)** |
| 3.A.8.L.2 | 281 (9.6%) | 1 (1.0%) |
| 3.B.1.I | 238 (8.1%) | 12 (11.4%) |
| 1.A.1.Z | 194 (6.6%) | 2 (1.9%) |
| 3.A.8.C | 177 (6.0%) | 7 (6.7%) |
| 3.A.8.A | 169 (5.7%) | 1 (1.0%) |
| 1.B | 168 (5.7%) | 13 (12.4%) |
| 3.A.8.Q | 108 (3.7%) | 0 (0.0%) |
| 3.A.8.I | 88 (3.0%) | 10 (9.5%) |
| 3.A.6.C | 87 (3.0%) | 1 (1.0%) |
| 3.B.1.B | 83 (2.8%) | 5 (4.8%) |
| **Top subgroups by wastewater isolate count** | **Clinical (%)** | **Wastewater (%)** |
| 1.B | 168 (5.7%) | 13 (12.4%) |
| 3.B.1.I | 238 (8.1%) | 12 (11.4%) |
| 3.A.8.N | 7 (0.2%) | 11 (10.5%) |
| 3.A.8.I | 88 (3.0%) | 10 (9.5%) |
| 3.A.8.C | 177 (6.0%) | 7 (6.7%) |
| 3.A.8.X | 45 (1.5%) | 7 (6.7%) |
| 3.B.1.B | 83 (2.8%) | 5 (4.8%) |
| 1.A.1.E | 26 (0.9%) | 5 (4.8%) |
| 3.A.21.B | 10 (0.3%) | 4 (3.8%) |
| 1.A.1.A | 55 (1.9%) | 3 (2.9%) |

After removing identical same-day wastewater isolates (zero SNP differences) to ensure robust comparative analysis, we identified 105 unique wastewater isolates. These wastewater-derived sequences demonstrated remarkable genomic concordance with the clinical population—90% matched clinical isolates within 3 SNPs and 48% showed exact matches (zero SNP differences) (**Supplementary Table 2**). These wastewater isolates mapped to 610 clinical isolates across 24 healthcare facilities, including all 18 facilities with the highest case burden, confirming broad representation of the regional outbreak. Detailed phylogenetic analysis of clinical and wastewater samples within Clade I revealed multiple distinct subclades harboring clinically relevant resistance mutations (**Supplementary Figure 3**). The *ERG11* Tyr132Phe mutation, conferring azole resistance, emerged consistently across evolving lineages, with particular prevalence in subclade 1.B (12.4% of wastewater isolates) (**Table 1**). While this high prevalence could represent repeated sampling of sustained shedding from one or more persistent colonization/case sources within the hospital, the lack of patient-level longitudinal data prevents definitive source attribution. Clinical isolates, in contrast, represent confirmed unique patient cases. Temporal analysis demonstrated the extraordinary predictive power of wastewater surveillance across several lineages. While subclade 1.A.3 appeared in wastewater 13 days before detection of the first clinical isolates (**Figure 4A, C**), the most striking early warnings came from Clade III isolates, which exhibited distinct evolutionary patterns characterized by *FKS1* mutations (**Supplementary Figure 4**). Most significantly, subclade 3.A.8.X (6.7% of wastewater isolates) (**Figures 4B, D, and Table 1**) was detected in wastewater nearly five months (143 days) before identification of the first clinical isolate—the longest advance warning period observed in our study. Similarly, subclade 3.A.8.Z appeared almost four months (112 days) before identification of the first clinical isolate (**Figure 4B, E**).

**Figure 4.**
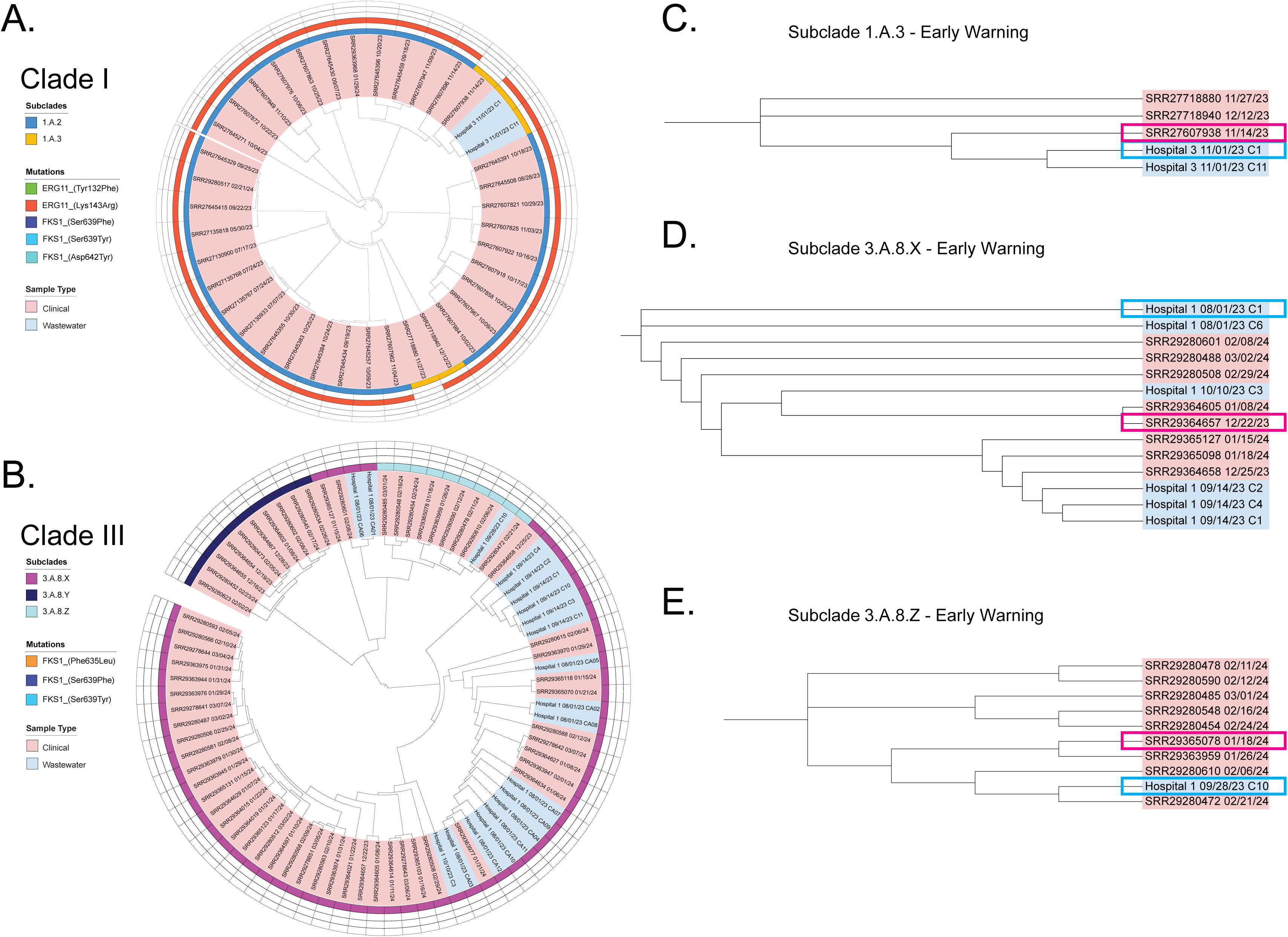
Phylogenetic analysis reveals early detection of resistance mutations in wastewater-derived *C. auris* isolates. **(A-B)** Maximum likelihood phylogenetic tree of Clade I and III isolates from Southern Nevada, showing major subclades and distribution of resistance mutations. Colored branches indicate distinct evolutionary lineages carrying different combinations of resistance-associated variants. (**C-E**) Detailed analysis of specific Clade I and III subclades showing early detection of subclades in wastewater: (**C**) Subclade 1.A.3, (**D**) Subclade 3.A.8.X, and (**E**) Subclade 3.A.8.Z. Red and blue boxes indicate clinical and wastewater isolates respectively and early detection of subclades in wastewater.

Comparative analysis revealed distinct resistance mutation patterns between sample types. *FKS1* mutations showed significantly higher prevalence in wastewater isolates (20.0%) compared to clinical isolates (2.1%), with wastewater variants dominated by Asp642Tyr (9 isolates), Ser639Phe (7 isolates), and Ser639Tyr (4 isolates) (**Table 2**). Mutations in *ERG11* maintained similar frequencies between environments (wastewater: 29.4%; clinical: 30.0%), predominantly Lys143Arg and Tyr132Phe mutations (**Table 3**). The *FUR1* mutation landscape also showed comparable rates (wastewater: 9.2%; clinical: 7.2%), though clinical isolates exhibited greater variant diversity (**Table 3**).

**Table 2.** FKS1 mutations in clinical and wastewater isolates.

| **FKS1 mutation comparisons** | <u>Clinical</u> | <u>Wastewater</u> |
| --- | --- | --- |
| no FKS1 mutation | 2913 | 88 |
| FKS1 (Asp642Tyr) | 21 | 9 |
| FKS1 (Leu640Val) | 11 | 0 |
| FKS1 (Arg641Ser) | 5 | 0 |
| FKS1 (Ser639Phe) | 5 | 7 |
| FKS1 (Ser639Tyr) | 5 | 4 |
| FKS1 (Arg1354His) | 3 | 0 |
| FKS1 (Leu638Phe) | 3 | 0 |
| FKS1 (Arg641Gly) | 2 | 0 |
| FKS1 (Phe635Leu) | 2 | 1 |
| FKS1 (Ile1361Thr) | 1 | 0 |
| FKS1 (Ile301Phe) | 1 | 0 |
| FKS1 (Leu686Phe) | 1 | 0 |
| FKS1 (Met690Ile) | 1 | 0 |
| FKS1 (Phe635del) | 1 | 0 |
| FKS1 (Phe635Tyr) | 1 | 0 |
| FKS1 (Ser1358Phe) | 1 | 0 |
| **FKS1 mutation summary** | <u>Clinical</u> | <u>Wastewater</u> |
| FKS1 mutation total | 64 | 21 |
| total auris | 2977 | 105 |
| % FKS1 mutation | 2.1% | 20.0% |

**Table 3.** ERG11 and FUR1 mutations in clinical and wastewater isolates.

| **ERG11 and FUR1 mutation comparisons** | <u>Clinical</u> | <u>Wastewater</u> |
| --- | --- | --- |
| no FUR1 or ERG11 mutation | 2019 | 77 |
| ERG11 (Lys143Arg) | 596 | 15 |
| ERG11 (Lys143Arg), FUR1 (Ala151Thr) | 1 | 0 |
| ERG11 (Lys143Arg), FUR1 (Ala88Val) | 1 | 0 |
| ERG11 (Lys143Arg), FUR1 (Arg101Leu) | 47 | 1 |
| ERG11 (Lys143Arg), FUR1 (Arg87Lys) | 4 | 0 |
| ERG11 (Lys143Arg), FUR1 (Gln16*) | 23 | 0 |
| ERG11 (Lys143Arg), FUR1 (Glu115*) | 6 | 0 |
| ERG11 (Lys143Arg), FUR1 (Glu64*) | 38 | 1 |
| ERG11 (Lys143Arg), FUR1 (Gly106Asp) | 16 | 0 |
| ERG11 (Lys143Arg), FUR1 (Gly176Asp) | 2 | 0 |
| ERG11 (Lys143Arg), FUR1 (Leu142fs) | 2 | 0 |
| ERG11 (Lys143Arg), FUR1 (Leu20*) | 1 | 0 |
| ERG11 (Lys143Arg), FUR1 (Pro185del) | 1 | 0 |
| ERG11 (Lys143Arg), FUR1 (Pro4fs) | 1 | 0 |
| ERG11 (Lys143Arg), FUR1 (stop_lost&splice_region) | 2 | 0 |
| ERG11 (Lys143Arg), FUR1 (Tyr216Asn) | 1 | 0 |
| ERG11 (Lys143Arg), FUR1 (Gly207Asp) | 0 | 2 |
| ERG11 (Lys143Arg), FUR1 (Val83Asp) | 4 | 0 |
| ERG11 (Tyr132Phe) | 148 | 7 |
| ERG11 (Tyr132Phe), FUR1 (Arg104*) | 0 | 6 |
| FUR1 (Ala88Thr) | 9 | 0 |
| FUR1 (Cys81Tyr) | 2 | 0 |
| FUR1 (Gly207Asp) | 3 | 0 |
| FUR1 (Gly35fs) | 4 | 0 |
| FUR1 (Val103Gly) | 46 | 0 |
| **ERG11 and FUR1 mutation summary** | <u>Clinical</u> | <u>Wastewater</u> |
| ERG11 mutation total | 894 | 32 |
| FUR1 mutation total | 214 | 10 |
| total auris | 2977 | 105 |
| % ERG11 mutation | 30.0% | 30.5% |
| % FUR1 mutation | 7.2% | 9.5% |

Importantly, we identified two novel subgroups (new1.1 and new3.1) exclusively in wastewater surveillance, demonstrating its potential for early detection of emerging lineages (**Supplementary Table 2**). Subsequent phylogenetic analysis revealed that new1.1 was the first detection of what would later be annotated as subclade 1.A.3. The consistent temporal advantage of wastewater detection—ranging from 13 days to 143 days (nearly five months) across various subclades and resistance mutations— combined with high genomic concordance between wastewater and clinical isolates (90% within 3 SNPs) establishes this approach as an effective early warning system for emerging *C. auris* variants.

### Emergence of pan-resistant *C. auris* traced through paired wastewater and clinical surveillance

Building upon our observation of high genomic concordance between wastewater and clinical isolates, we next investigated whether culture-based wastewater surveillance could serve as an early indicator of emerging antifungal resistance. We conducted antifungal susceptibility testing (AST) across three major drug classes—azoles, echinocandins, and polyenes—using paired wastewater and clinical isolates (**Figure 5A-C**). To establish baseline resistance profiles, we first analyzed reference strains: CDC_CAU-09 (Clade I) exhibited high fluconazole resistance (≥256 mg/L) while maintaining echinocandin susceptibility (anidulafungin 0.19-0.25 mg/L, caspofungin 0.125-0.19 mg/L, micafungin 0.094-0.125 mg/L). In contrast, CDC_CAU-27 (Clade III) demonstrated broad drug susceptibility, with consistently low echinocandin MICs (0.094-0.19 mg/L) and moderate amphotericin B tolerance (0.75-1 mg/L) (**Figure 5B**).

**Figure 5.**
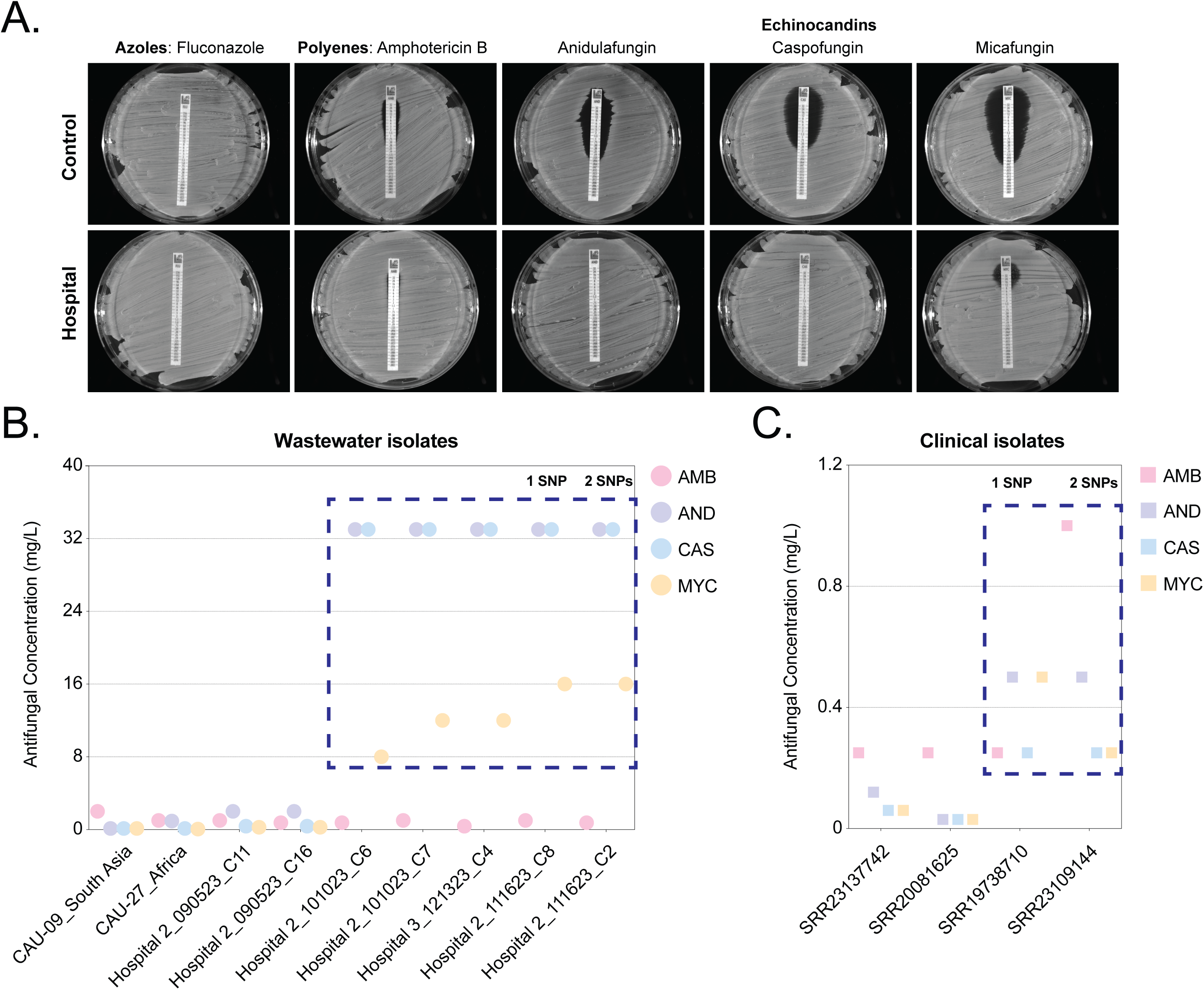
Temporal evolution of antifungal resistance profiles in *C. auris* isolates. **(A)** Representative antifungal susceptibility testing (AST) profiles showing drug responses against multiple antifungal classes for **(top)** reference strain CDC_CAU-09 (Clade I) and **(bottom)** a resistant Hospital 2 wastewater isolate showing elevated MICs. **(B)** Temporal progression of resistance in wastewater isolates from Hospital 2 and Hospital 3 (September-December 2023). Wastewater isolates are designated by collection location, date (MMDDYY), and isolate number (e.g., Hospital 2_090523_C11 indicates Hospital 2, September 5, 2023, isolate C11). Early isolates (September 2023) show baseline resistance patterns, evolving to echinocandin resistance (≥32 mg/L) in later isolates while maintaining amphotericin B susceptibility (0.50-1 mg/L). Blue dotted boxes highlight clinical isolates showing elevated MICs. **(C)** Phenotypic concordance between wastewater and clinical isolates. Blue dotted boxes highlight clinical isolates showing elevated MICs. Two key paired examples demonstrate the genetic-phenotypic relationships: Hospital 2_111623_C8 is separated by 1 SNP from clinical isolate SRR19738710, while Hospital 2_111623_C2 is separated by 2 SNPs from clinical isolate SRR23109144. Both pairs show similar resistance profiles, particularly for echinocandins and amphotericin B susceptibility patterns. Legend: Amphotericin B (AMB), Anidulafungin (AND), Caspofungin (CAS), and Micafungin (MYC).

Longitudinal AST profiling of select wastewater isolates revealed a striking stepwise evolution of drug resistance (**Figure 5B**). The first signs of emerging echinocandin resistance appeared in Hospital 2 during early October 2023. While these isolates were predominantly Clade III, the phenotypic resistance patterns showed more complex relationships with genomic changes than simple one-to-one correspondence with known resistance mutations. Specifically, isolates C6 and C7 developed high-level resistance to echinocandins (anidulafungin and caspofungin ≥32 mg/L, micafungin 8-12 mg/L). This resistance pattern subsequently spread, with isolates from both Hospital 2 and Hospital 3 (C4, C8, C2) exhibiting echinocandin resistance (≥32 mg/L) by November-December 2023. Throughout this progression, amphotericin B (AMB) susceptibility remained remarkably stable (0.50-1 mg/L), suggesting highly specific selective pressure targeting echinocandin resistance mechanisms.

To validate the predictive potential of wastewater surveillance for resistance emergence, we performed paired AST analysis comparing wastewater isolates with their closest clinical matches. These pairs were identified through SNP-based phylogenetic analysis (**Figure 5C**)^24,25^. To systematically evaluate phenotypic concordance, we first identified wastewater-clinical isolate pairs based on genomic similarity, focusing on those separated by less than two SNPs. This analysis revealed robust phenotypic concordance between paired isolates. For example, wastewater isolate Hospital 2_111623_C8 (separated by one SNP from clinical isolate SRR19738710) showed similar higher-level echinocandin resistance (MIC ≥32 mg/L). Similarly, Hospital 2_111623_C2 and its clinical counterpart SRR23109144 (two SNPs apart) both exhibited parallel resistance patterns across multiple drug classes. The strong correlation between resistance profiles in wastewater and clinical isolates supports wastewater surveillance as a relevant early warning system for emerging antifungal resistance. However, AST remains essential for definitively characterizing resistance patterns in newly detected isolates to enable truly real-time monitoring.

### Transcriptomic Analysis Reveals Novel Stress Adaptation Mechanisms in *C. auris*

To understand the molecular mechanisms enabling *C. auris* adaptation to environmental and therapeutic stresses, we performed comprehensive transcriptomic profiling across multiple physiologically relevant conditions (**Figure 6A**). We simultaneously exposed wastewater isolates to thermal stress (37°C and 42°C), antifungal pressure (fluconazole), and antibiotic exposure, generating high-depth RNA sequencing data (5-10 million reads per sample) to characterize stress-response transcriptomes.

**Figure 6.**
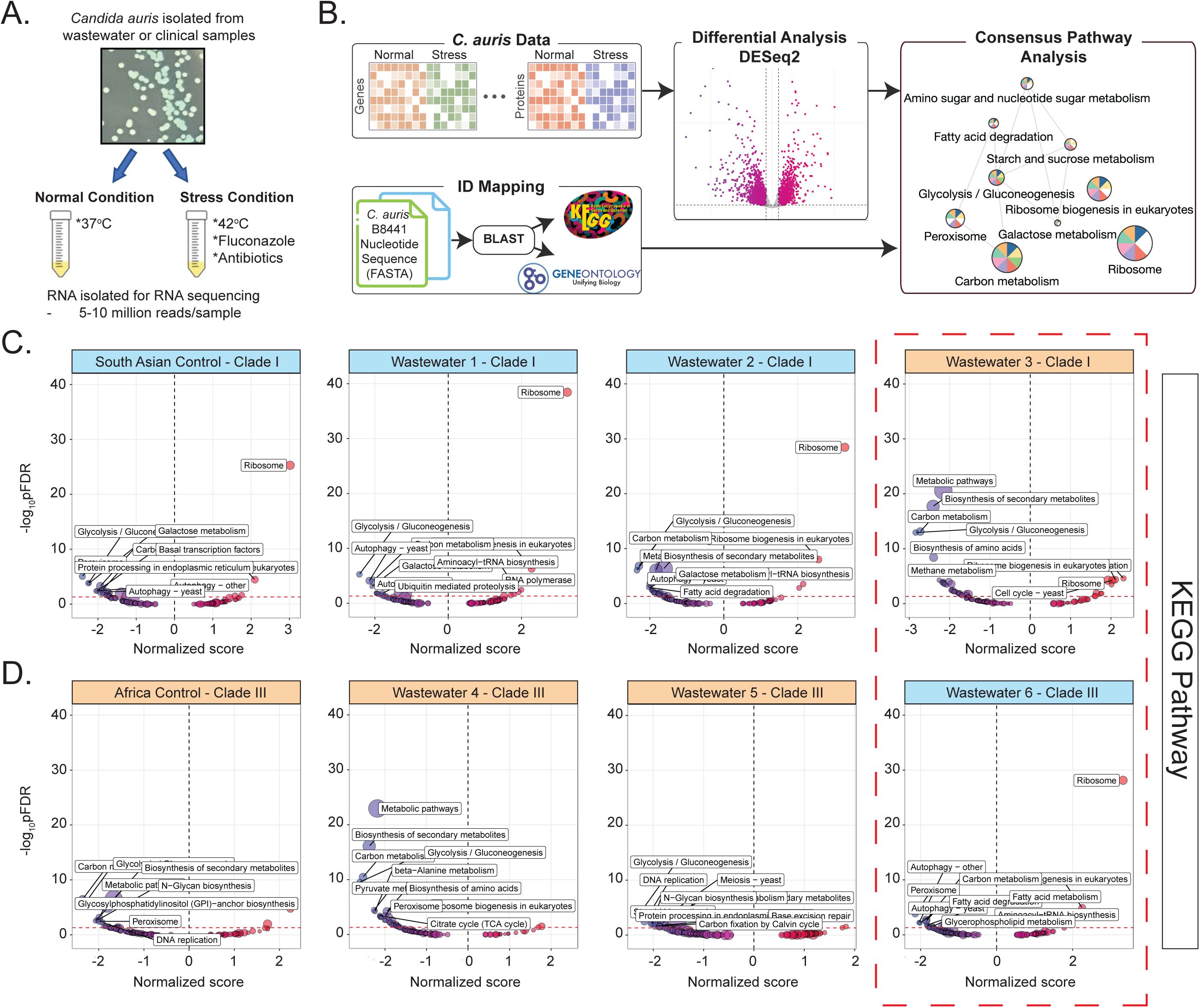
Comparative transcriptional profiling reveals distinct stress response mechanisms in clinical and environmental *C. auris* isolates. **(A)** Experimental design for stress response analysis. Wastewater and clinical *C. auris* isolates were exposed to multiple stressors (37°C, 42°C, fluconazole, and antibiotics) with paired normal condition controls. RNA sequencing generated >5 million reads per sample. **(B)** Bioinformatic workflow for consensus pathway analysis. RNA-seq data was processed through alignment to *C. auris* B8441 reference genome, followed by gene identification using BLAST, differential expression analysis using DESeq2, and pathway enrichment analysis using CPA. **(C)** KEGG pathway enrichment analysis of Clade I isolates. Plots show normalized pathway scores (-log_10_ pFDR vs. normalized enrichment score) for Clade I control strain and three wastewater isolates. While wastewater isolates 1 and 2 maintain similar pathway activation patterns to the control (dominated by changes in ribosome biogenesis and carbon metabolism), wastewater isolate 3 shows distinct pathway utilization focused on DNA replication and cell cycle processes. **(D)** KEGG pathway analysis of Clade III isolates comparing Clade III control strain with three wastewater isolates. Strikingly, wastewater isolate 6 exhibits opposite pathway regulation compared to other samples, with significant downregulation of peroxisome and fatty acid degradation pathways and unique activation of ribosome biogenesis.

We applied our consensus pathway analysis (CPA)^28,29^ pipeline to systematically identify stress-activated pathways in *C. auris* (**Figure 6B**). This computational framework integrated *de novo* transcriptome assembly with mapping to the *C. auris* B8441 reference genome, followed by BLAST-based gene annotation and pathway enrichment analysis. Differential expression analysis using DESeq2 revealed distinct stress-specific transcriptional signatures that we mapped to cellular pathways using stringent statistical criteria.

Analysis of Clade I isolates revealed a hierarchical organization of stress responses (**Figure 6C**). The reference strain exhibited a canonical stress response, characterized by coordinated upregulation of ribosomal pathways (normalized enrichment score 3.1), ribosome biogenesis (normalized enrichment score 2.1), and carbon metabolism (normalized enrichment score -1.9). Wastewater isolates 1 and 2 maintained this core response while activating additional survival mechanisms, including aminoacyl-tRNA biosynthesis pathways, suggesting enhanced protein quality control. Interestingly, wastewater isolate 3 displayed a fundamentally altered stress response, characterized by metabolic pathway deactivation and upregulation of cell cycle regulation, with absence of traditional stress responses.

Clade III isolates demonstrated parallel evolutionary adaptations (**Figure 6D**). While the reference strain and wastewater isolates 4 and 5 showed balanced activation of primary metabolic pathways, wastewater isolate 6 revealed complete rewiring of cellular stress responses. This isolate downregulated peroxisome function and fatty acid metabolism while strongly activating ribosomal pathways. It uniquely suppressed efferocytosis pathways and demonstrated fundamental restructuring of carbon metabolism through beta-alanine and propanoate pathways, indicating novel stress adaptation mechanisms.

These transcriptional profiles provide unprecedented insights into *C. auris* stress adaptation, demonstrating that antifungal resistance emerges through complex reprogramming of cellular pathways rather than simple target modification. The extensive metabolic rewiring observed in wastewater isolates 3 and 6 suggests the evolution of adaptive mechanisms involving dynamic regulatory shifts in stress response networks, energy metabolism, and cell wall remodeling pathways. These changes likely enhance cellular resilience, contributing to stable resistant phenotypes that persist in clinical settings. Furthermore, the identification of differentially expressed genes linked to oxidative stress resistance, membrane integrity, and metabolic plasticity reveals previously uncharacterized vulnerabilities that could serve as novel therapeutic targets for combating this highly adaptable fungal pathogen.

## Discussion

Our integrated analysis of *Candida auris* using wastewater-based epidemiology (WBE) establishes a new paradigm for tracking fungal pathogen evolution and resistance emergence. The demonstration of up to 5 months advance detection of subclades compared to clinical surveillance, combined with 90% genomic concordance between wastewater and clinical samples, reinforces how WBE can be used as a powerful tool for proactive resistance surveillance. These findings provide compelling evidence for incorporating environmental/wastewater monitoring into global strategies addressing the rising threat of emerging pathogens and drug resistance^22,30^. The early detection of resistance mutations, including *FKS1* Phe635Leu and *ERG11*/*FKS1* variant combinations, provides a critical window for intervention before resistant strains establish dominance in healthcare settings. This advance warning enables implementation of targeted control measures, including enhanced screening, contact precautions, and antifungal stewardship^31^. However, the marked difference in *FKS1* mutation prevalence between wastewater (20.0%) and clinical isolates (2.1%) highlights the complex relationship between environmental detection and clinical outcomes, emphasizing the need for careful interpretation of wastewater surveillance data.

To better understand these wastewater findings, our longitudinal analysis reveals key insights into the stepwise evolution of echinocandin resistance. The progression from elevated anidulafungin MICs to pan-echinocandin resistance, while maintaining amphotericin B susceptibility, suggests evolutionary constraints on simultaneous resistance development^32^. These patterns likely reflect specific selective pressures within healthcare environments^33^, possibly driven by antimicrobial usage patterns or unique wastewater system dynamics. Understanding these evolutionary constraints could inform therapeutic strategies that extend the efficacy of existing antifungal agents. Complementing these findings, our transcriptomic analyses uncover previously unrecognized dimensions of *C. auris* adaptation. The extensive metabolic rewiring observed in resistant isolates, including downregulation of peroxisome and fatty acid degradation pathways^34^, challenges traditional resistance paradigms focused solely on drug target modifications. The engagement of novel pathways, particularly in carbon metabolism and cellular stress responses, reveals fundamental adaptation mechanisms that could serve as innovative therapeutic targets.

Despite these promising results, implementation of WBE faces several practical challenges that warrant attention. These include standardizing sampling protocols across diverse healthcare settings, understanding facility-specific influences on resistance patterns, and the inherent limitations of culture-based detection methods, as some cultures remain difficult to grow or isolate from complex wastewater matrices. This could create potential blind spots that may affect clinical decision-making. PCR-based detection methods may show cross-reactivity with other *Candida* species and require careful validation. Our findings raise important questions for future investigation: What drives the rapid emergence/evolution of resistance within healthcare facilities? How do novel metabolic adaptations influence pathogen fitness and transmission? Can this surveillance approach be extended to other microbial pathogens, including both fungi and bacteria? Addressing these questions requires integrating molecular biology, environmental science, and clinical epidemiology. The early detection of resistance variants, coupled with insights into previously undescribed adaptation mechanisms, demonstrates WBE’s value as an important tool for pathogen surveillance. Importantly, the identification of novel subclades and resistance mutations exclusively in wastewater highlights its unique value for uncovering cryptic resistance reservoirs.

Taken together, our study validates upstream (i.e., facility-specific) wastewater surveillance as a powerful approach for monitoring fungal pathogen evolution, providing a robust framework for integrating environmental and clinical data into comprehensive public health strategies. By advancing our understanding of *C. auris* resistance and adaptation mechanisms, we establish a foundation for proactive, scalable approaches to managing fungal outbreaks and combating the global challenge of antifungal resistance.

## Methods

### Wastewater sample collection, processing, and PCR analysis

Raw wastewater samples were collected weekly from three major hospitals and their corresponding WWTPs in Southern Nevada between August and December 2023. Similar to our previous study^35^, composite samples were placed on ice in the field and stored under refrigeration until processing (hold time < 36 h). For each sampling event, approximately 100 mL of wastewater was pelleted by centrifugation at 4,000 rpm for 10 minutes at 4°C. Total DNA was extracted from the pelleted solids using the DNeasy PowerSoil Pro Kit (Qiagen Catalog #47016). The pellet was resuspended in 800 µL of Solution CD1 and transferred to the PowerBead Pro tube. Nucleic acid extraction was then carried out according to the manufacturer’s instructions. Hospital-specific sampling points were established within each facility’s sewer network to capture wastewater flows before mixing with other municipal sewage streams^23,30^. At WWTPs, grab (due to practical limitations at Facility 4A) influent samples were collected^36,37^. This sampling strategy enabled comparison between facility-specific (i.e., hospital) signals and the more dilute pathogen concentrations typically observed at downstream treatment plants. Sample processing was conducted in dedicated biosafety facilities following standard protocols for handling potentially infectious materials. All sample collection points were georeferenced and sample metadata (time, date, location, flow conditions) were recorded to maintain data quality and facilitate spatial-temporal analyses.

*C. auris* DNA in the samples was quantified by qPCR based on a CDC assay that targets the internal transcribed spacer 2 (ITS2) gene^16^. The sequences for the primers are: 1) For: 5’- CAG ACG TGA ATC ATC GAA TCT-3’, and 2) Rev: 5’- TTT CGT GCA AGC TGT AAT TT-3’. The Probe sequence is: 5’-FAM-AAT CTT CGC GGT GGC GTT GCA TTC A-BHQ1-3’. A synthetic DNA gBlock Gene Fragment qPCR standard for *C. auris* was utilized at different concentrations to create a standard curve, as shown previously for other wastewater targets^22^. Primers, probes, and standards were acquired from Integrated DNA Technologies (IDT, Coralville, IA). Nucleic acid samples and standards were analyzed by qPCR using the Bio-Rad CFX Opus 96 instrument with the Millipore Sigma KiCqStart Probe qPCR ReadyMix (Catalog #KCQS04) as the reaction master mix. The final quantity in gene copies (gc) was divided by the equivalent sample volume (ESV), which was constant at 2 mL; after unit conversion, this yielded a final *C. auris* DNA concentration in gc/L which represents the solids-associated signal.

### Wastewater amplicon panel, library preparation, and bioinformatics

We designed an amplicon panel to detect antifungal resistance-associated genes in *C. auris*. The panel targeted eleven genes implicated in resistance mechanisms: *CDR1*, *ERG3*, *ERG6*, *ERG11*, *FKS1*, *HSP90*, *MEC3*, *MLH1*, *MRR1A*, *TAC1B*, and *UPC2*. The panel were designed using the *C. auris* strain B8441 V2 reference genome (GCA_002759435.2). The first panel, generated using PrimalScheme version 1.4.1^38^, comprised 345 primer pairs. The second panel, developed using Olivar version 1.1.1^39^, contained 435 primer pairs and incorporated the Simulated Annealing Design using Dimer Likelihood Estimation (SADDLE) algorithm^40^ to minimize primer dimer formation and non-specific amplification. To enable species-level discrimination, we identified unique sequence markers for both *C. auris* and *C. albicans* using unikmer version 0.19.2 and incorporated these sequences into the panel.

The primer panel was synthesized as oPools Oligo Pools by IDT. Library preparation consisted of two main steps. First, we performed targeted amplification using CleanPlex mPCR Mix (Paragon Genomics, Catalog #219011) with our custom primer pools. Second, we processed the amplified products using the NEBNext® Ultra™ II DNA Library Prep Kit for Illumina (Catalog #E7645L) and NEBNext® Multiplex Oligos for Illumina (Catalog #E7335L) from New England Biolabs. The final multiplexed libraries were sequenced on an Illumina NextSeq 1000 platform using a NextSeq 1000/2000 P1 flow cell with 300 cycles.

Raw sequencing reads were processed using fastp v0.23.4 to remove Illumina adapters, filtering low quality reads (Phred quality score <20), and trimming of polyG tails^41^. The trimmed reads were aligned to a custom reference genome custom reference genome consisting of the concatenated reference FASTA referenced earlier using BWA MEM v0.7.17-r1188 with default parameters^42^. Post-alignment sorting and indexing was performed using samtools v1.15.1^43^. Amplicon primers were trimmed using fgbio TrimPrimers v2.4.0 and fgbio FilterBam was used to trim reads shorter than 40 base pairs. Variant calling was performed using ivar v1.4.3^44^. iVar’s TSV output was converted to VCF for annotation with SnpEff, using a GFF file generated from the concatenated reference^45^. Samples passing the following QC metrics were retained for downstream analysis: at least 50% of positions covered at ≥100x depth and a minimum read depth of 100 per SNP to balance sensitivity and specificity in variant detection.

### Whole genome sequencing of wastewater and clinical samples and bioinformatics

We performed comprehensive genomic analysis of *C. auris* isolates from both clinical sources (collected during 2021-2024) and wastewater using two pipelines: the MycoSNP nextflow pipeline version 1.5^46,47^ and TheiaEuk fungal genome specific workflow^24^. MycoSNP was used for reference-based alignment, variant calling, and downstream population-level analyses, while TheiaEuk provided species classification, subclade (clade) typing, and SNP-sharing analyses. For reference-based alignment, we used clade-specific genomes: *C. auris* B11205 (GenBank assembly GCA_016772135.1) for Clade I isolates and *C. auris* B11211 (GenBank assembly GCA_002775015.1) for Clade III isolates. To minimize false-positive variant calls, we first identified and masked repetitive elements (approximately 2% of the genome) using MUMmer (version 4.0) nucmer followed by Bedtools makefasta^48,49^. Raw sequencing reads underwent quality control and preprocessing using FaQCs version 2.10 before alignment to the appropriate reference genome using BWA MEM version 0.7.17-r1188^42,50^. We processed the resulting alignments using Samtools version 1.15 to convert and sort sequence alignment map (SAM) files to binary alignment map (BAM) format^43^. Further refinement of alignments included marking duplicate reads with Picard version 2.26.10 MarkDuplicates and soft-clipping unaligned read regions using Picard CleanSam. For variant calling, we employed GATK HaplotypeCaller version 4.2.5.0, configured for haploid genome analysis. We applied stringent filtering criteria: quality by depth > 2.0, Fisher strand bias < 60.0, mapping quality > 40.0, and read depth > 30^51^. Additional quality control measures included filtering for minimum genotype quality (GQ) ≥ 50 and alternative allele frequency ≥ 80% in allele depth. Samples were kept for further analysis if they passed all quality control filters, including the requirement that they exhibit at least 70 percent average coverage across *C. auris* B8441V2 scaffolds 1 - 10 and 12 - 14 at a minimum of 30× depth. Variant annotation was performed using SnpEff and SnpSift version 5.2, with results exported to tab-separated format for downstream analysis^45,52^. To enable population-level analyses, we merged individual variant calls using GATK CombineGVCFs. We identified high-confidence SNPs showing ≤ 10% ambiguity across the sample set and concatenated them into a multi-fasta alignment using custom Python scripts^53^. Finally, we calculated SNP distance matrices using snp-dists and constructed maximum likelihood phylogenetic trees using RAxML-NG. Tree visualization and annotation were performed using Interactive Tree Of Life^54^.

### MALDI-TOF mass spectrometry

Wastewater samples were concentrated by centrifugation and approximately 10 mg of pelleted material was resuspended in 300 µL HPLC-grade deionized water. Cellular proteins were extracted using a modified ethanol-formic acid protocol. Briefly, samples were mixed with 900 µL pure ethanol, centrifuged (17,000 × *g*, 2 min, 25°C), and the resulting pellet was washed with an additional ethanol step to ensure complete removal of contaminants. After air-drying, proteins were extracted using 40 µL of 70% formic acid followed by freeze-thaw treatment (−80°C for 20 min, thawed 10 min at 25°C). The extract was mixed with an equal volume of acetonitrile, centrifuged (17,000 × *g*, 2 min, 25°C), and 70 µL of supernatant was collected. For MALDI-TOF MS analysis, 0.5 µL of protein extract was spotted in triplicate onto a MALDI target plate and overlaid with 0.5 µL of matrix solution (α-cyano-4-hydroxycinnamic acid, 100 mg/mL in 50% acetonitrile, 2.5% trifluoroacetic acid). Mass spectra were acquired using a Shimadzu MALDI-8020 mass spectrometer operated in positive linear mode (mass range 2,000-20,000 Da). The instrument was calibrated using DH5α competent cells (Thermo Fisher) as an external standard. All samples were analyzed in technical triplicate and representative spectra are shown.

### Antifungal susceptibility tests

Minimum inhibitory concentrations (MICs) were determined using the agar diffusion method with commercially available MIC test strips (Liofilchem). Standardized inocula were prepared by adjusting yeast suspensions to 2.5 × 10⁶ cells/mL in YPD broth (Sigma Catalog #Y1375). YPD agar plates (MilliporeSigma) were inoculated using standardized triple-directional streaking technique to ensure uniform cell distribution. MIC test strips containing antifungal agents were applied to the inoculated plates and incubated at 37 ± 1°C for 18-20 h under aerobic conditions. MIC values were determined at the intersection of the inhibition ellipse with the test strip gradient markings. Discrete microcolonies within the inhibition zones were excluded from MIC determinations as per standard interpretative criteria. All susceptibility tests were performed in biological triplicate with appropriate quality control strains (*C. auris* CAU-09 and CAU-27).

### RNA sequencing and data analysis

*C. auris* isolates were grown in YPD (Sigma Catalog #Y1375, 37°C, overnight) or SSDB (Thomas Scientific Catalog #CHM01P620, 42°C, 48 h). Cells were pelleted (2,900 × *g*, 5 min), resuspended in YR Digestion Buffer (Zymo Catalog #R1001-1) with 25 U Zymolyase (Zymo E1004), and incubated at 37°C for 60 min. Additional YR buffer and equal volume of ethanal (95-100%) were added and RNA was purified according to the manufacturer’s instructions for the RNeasy PowerMicrobiome Kit (Qiagen Catalog #26000-50), including DNase I treatment and elution in 54 µL RNase-free water. Libraries were prepared using the Illumina Stranded mRNA Prep, Ligation Kit (Illumina Catalog #20040534) with IDT RNA UD Indexes Set A (Illumina Catalog #20040553). The library preparation followed the manufacture’s protocol. Briefly, RNA (1 µg) was incubated with RNA Purification Beads (RPBX) to capture mRNA, washed, eluted, and fragmented using Elute, Prime, Fragment High Mix (EPH3). First- and second-strand cDNA synthesis used First Strand Synthesis Mix (FSA + RVT) and Second Strand Marking Master Mix (SMM), followed by 1.8X bead cleanup with NucleoMag NGS Clean-up beads (Macherey Nagel Catalog #744970.500). Adenylation was performed with A-Tailing Mix (ATL4), followed by adapter ligation with RNA Index Anchors and Ligation Mix (LIGX), and cleanup with a 0.8X bead ratio. Amplification was performed using Enhanced PCR Mix (EPM) and UD Index primers (14 cycles). Final libraries were cleaned with 1X beads and eluted in 17 µL RSB. Library quality was verified using the TapeStation system with High Sensitivity D1000 ScreenTape (Agilent Catalog #5067-5584), and concentrations were measured using the Qubit Flex Fluorometer with Qubit 1x dsDNA HS Assay Kit (Thermo Fisher Catalog #Q33231). Libraries were diluted to 2 nM, pooled, and sequenced on the NextSeq 1000 system using NextSeq 1000/2000 P2 Reagents (300 Cycles) v3 kit (Illumina Catalog #20046813).

We analyzed transcriptional profiles of wastewater-derived *C. auris* isolates using the nf-core/rnaseq pipeline^47^. All analyses were performed using the *C. auris* B8441 reference genome (version s01-m03-r13) from the Candida Genome Database^55^. Quality control and read preprocessing involved multiple steps to ensure data integrity. First, we performed adapter removal and quality trimming using Trim Galore! version 0.6.7 with Cutadapt version 3.4, applying stringent quality score thresholds. To eliminate potential contamination, we screened reads using BBTools BBSplit version 38.90 for genomic DNA and SortMeRNA version 4.3.6 for ribosomal RNA sequences. For transcriptome analysis, we aligned processed reads to the reference genome using STAR aligner version 2.7.9a, followed by transcript quantification with Salmon version 1.10.1^56^. The resulting alignments were sorted and indexed using Samtools version 1.17, with duplicate reads marked using Picard version 2.26.10 MarkDuplicates. We then assembled and quantified transcripts using StringTie version 2.2.1 to generate comprehensive count matrices^57^. To validate transcriptome-derived genetic variants, we performed variant calling on RNA-seq alignments using Freebayes version 1.3.6. We applied stringent filtering criteria (allele frequency ≥ 0.9, sequencing depth ≥ 10) and compared these variants with our whole-genome sequencing results to confirm genotype consistency across different sequencing approaches. Differential expression analysis was conducted using the nf-core/differentialabundance pipeline with DESeq2 version 1.34.0. We identified significantly differentially expressed genes using threshold criteria of adjusted p-value ≤ 0.05 and absolute log_2_ fold change ≥ 1. This analysis revealed distinct transcriptional signatures associated with stress responses and antifungal resistance, as detailed in our results section.

### Consensus Pathway Analysis (CPA) data analysis

To examine the biological pathways affected under different stress conditions, we developed a comprehensive pathway analysis framework integrating both KEGG (release 111.0)^58^ and Gene Ontology (GO, release 2024-09-08)^59^ annotations. Given that neither database directly supported Candida Genome Database identifiers, we first established robust gene mapping protocols. For KEGG pathway mapping, we obtained nucleotide sequences directly from the KEGG database. GO gene sequences were sourced from the NCBI *C. auris* assembly ASM301371v2. We then mapped *C. auris* genes to their KEGG and GO counterparts using BLAST (version 2.5.0)^60,61^, implementing a stringent similarity threshold of 95% to ensure high-confidence annotations. Pathway enrichment analysis was performed using our previously developed Consensus Pathway Analysis (CPA)^29,62^ framework. We employed the *runGeneSetAnalysis()* function from the RCPA package (version 0.2.2) to execute Fast Gene Set Enrichment Analysis (FGSEA)^63^ on differential expression results from DESeq2. Statistical significance was assessed using Benjamini-Hochberg False Discovery Rate^64^ correction for multiple testing. To visualize pathway enrichment patterns, we generated volcano plots comparing normalized enrichment scores against -log_10_ adjusted p-values, revealing distinct stress response signatures across different experimental conditions.

### Data sharing

Any request to access relevant data in this study should be sent to the corresponding authors.

## Declaration of interests

All authors declare no competing interests.

## Data Availability

All data produced in the present work are contained in the manuscript.

https://github.com/M-Moshi/UNLV-NPM-Candida-auris/

## Acknowledgments

ECO and VV are supported by NIH grant: MH109706. DG, CL, VV, and ECO are supported by Grant Number NH75OT000057-01-00 from the Centers for Disease Control and Prevention (CDC). TN, QHN, and HN are supported by NIH: R44GM152152 and U01CA274573. The project contents are solely the responsibility of the authors and do not necessarily represent the official views of the CDC. This work would have not been possible without the participation of the collaborating wastewater agencies in Southern Nevada. The corresponding authors have full access to all the data in the study and take responsibility for the integrity of the data and the accuracy of the data analysis.

---

**Supplementary Figure 1.**
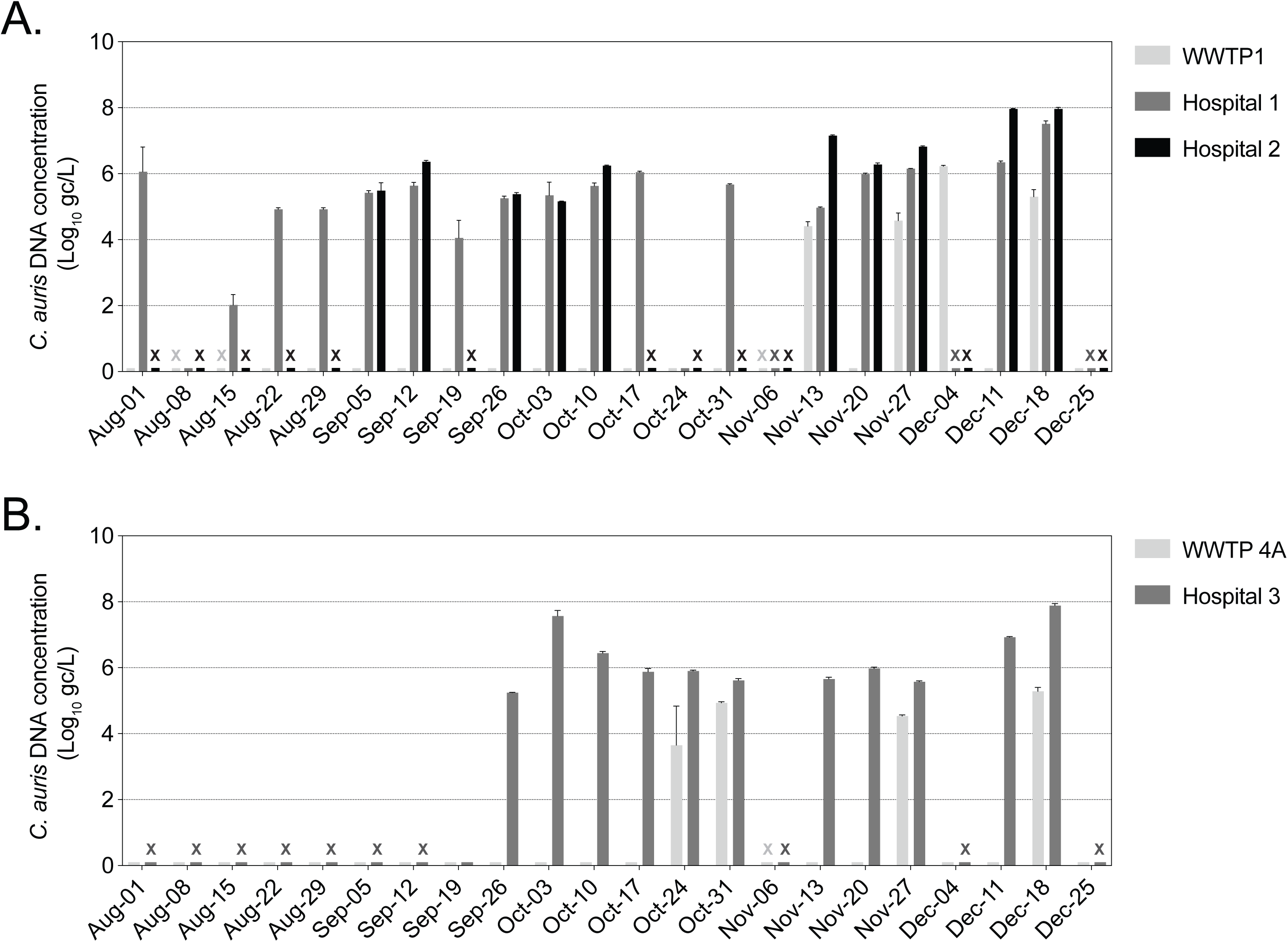

**Supplementary Figure 2.**
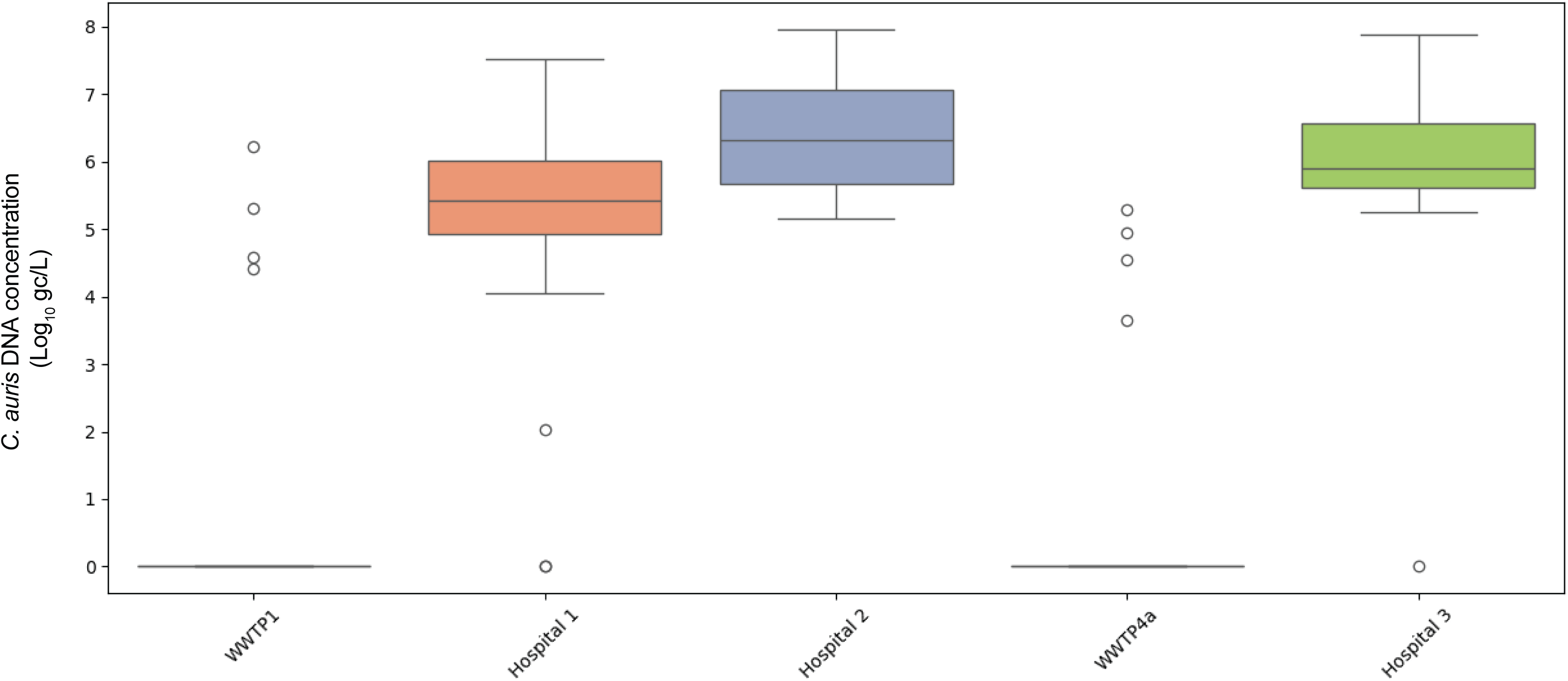

**Supplementary Figure 3.**
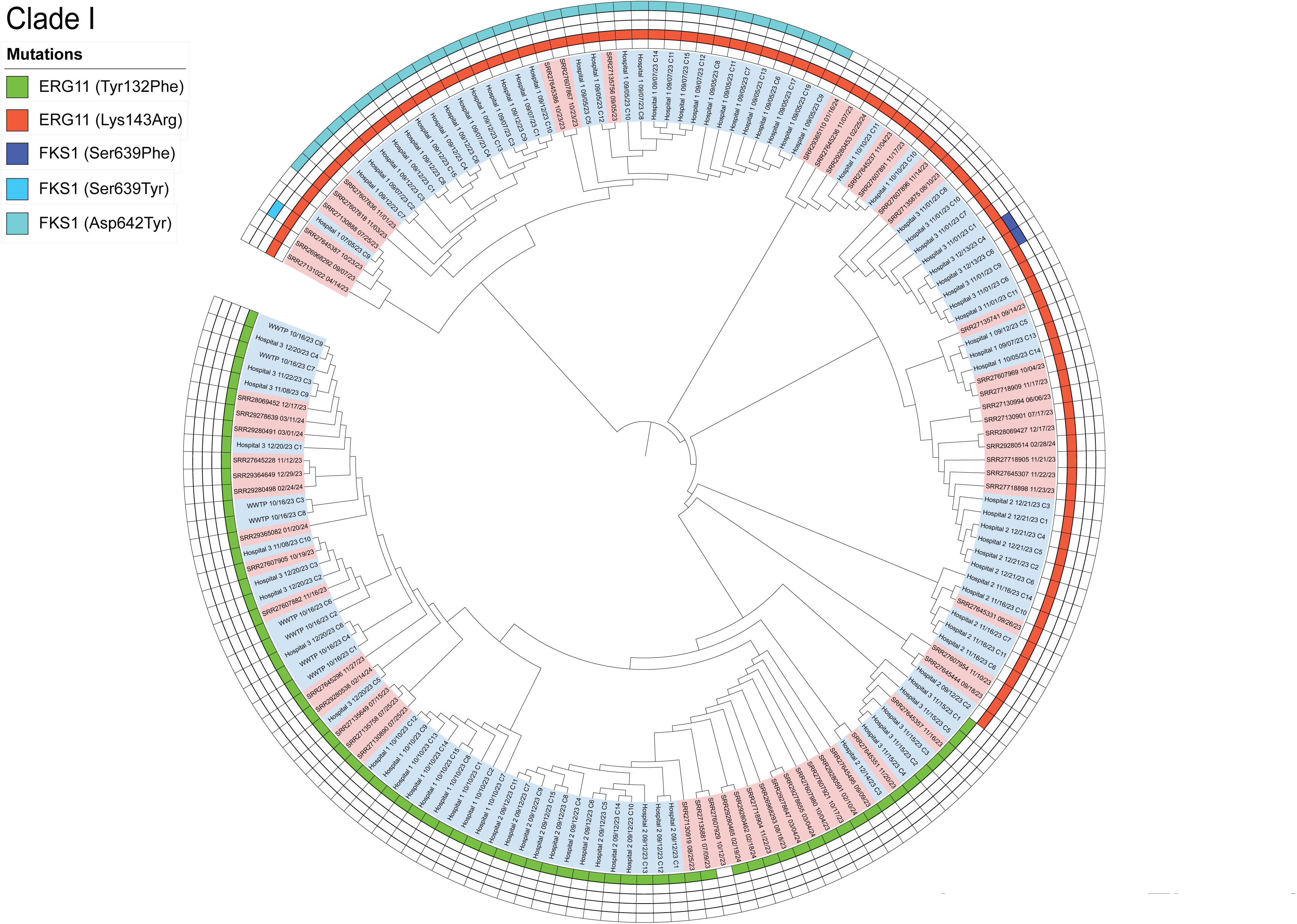

**Supplementary Figure 4.**
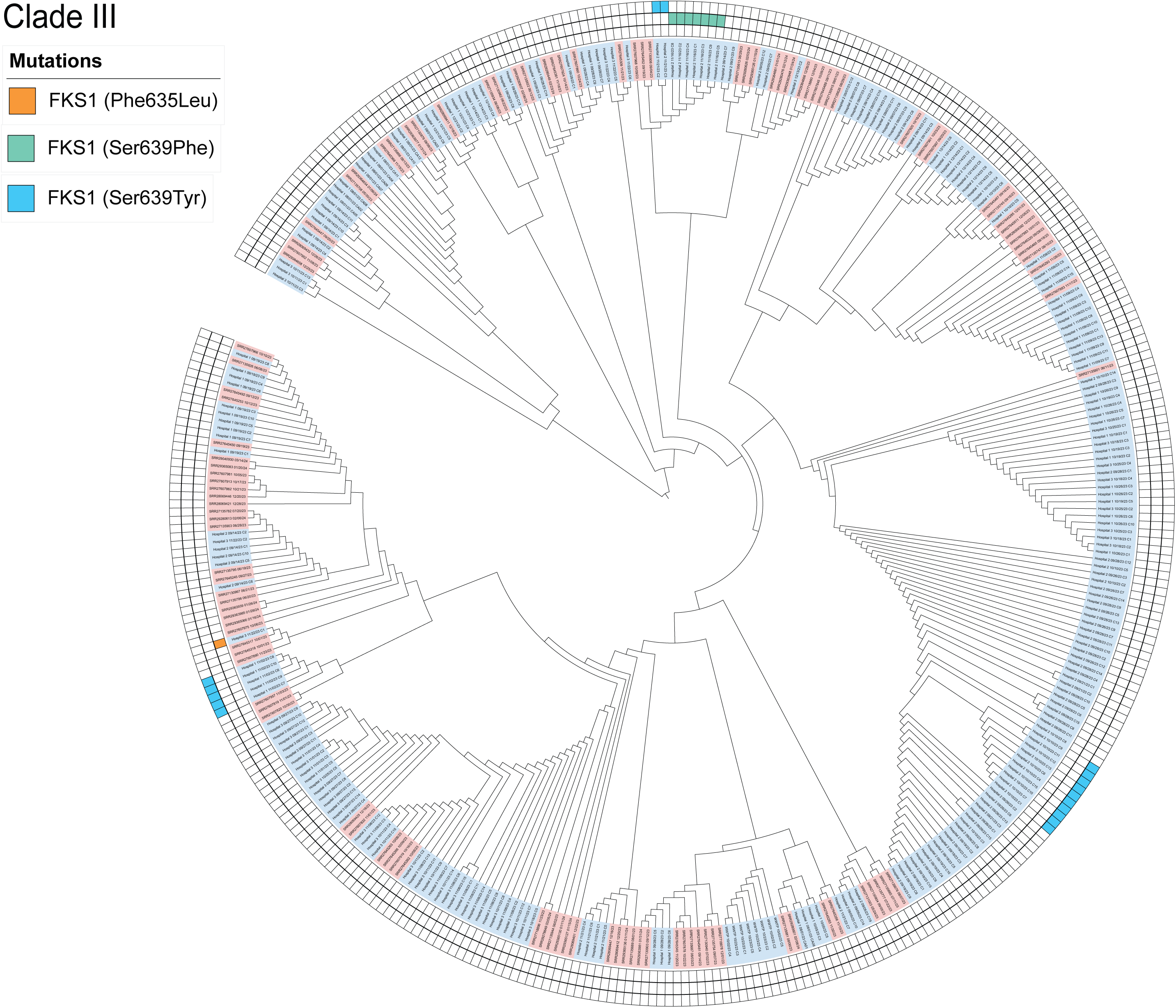

**Supplementary table 1.** Amplicon panel targeting 11 *C. auris* genes.

| Gene | Accession | Position: Start | Position: End |
| --- | --- | --- | --- |
| *CDR1* | PEKT02000001.1 | 348521 | 353047 |
| *C. albicans* unique | NC_032089.1 | 66764 | 67698 |
| *C. auris* unique | PEKT02000007.1 | 172646 | 173634 |
| *ERG11* | PEKT02000003.1 | 832733 | 834307 |
| *ERG3* | PEKT02000007.1 | 2369962 | 2371053 |
| *ERG6* | PEKT02000010.1 | 960572 | 961699 |
| *FKS1* | PEKT02000002.1 | 1002873 | 1008539 |
| *HSP90* | PEKT02000010.1 | 61761 | 63887 |
| *MEC3* | PEKT02000007.1 | 1041428 | 1042390 |
| *MLH1* | PEKT02000008.1 | 783713 | 785689 |
| *MRR1A* | PEKT02000007.1 | 3093802 | 3096522 |
| *TAC1B* | PEKT02000009.1 | 814591 | 817197 |
| *UPC2* | PEKT02000001.1 | 559186 | 561039 |

**Supplementary table 2.**
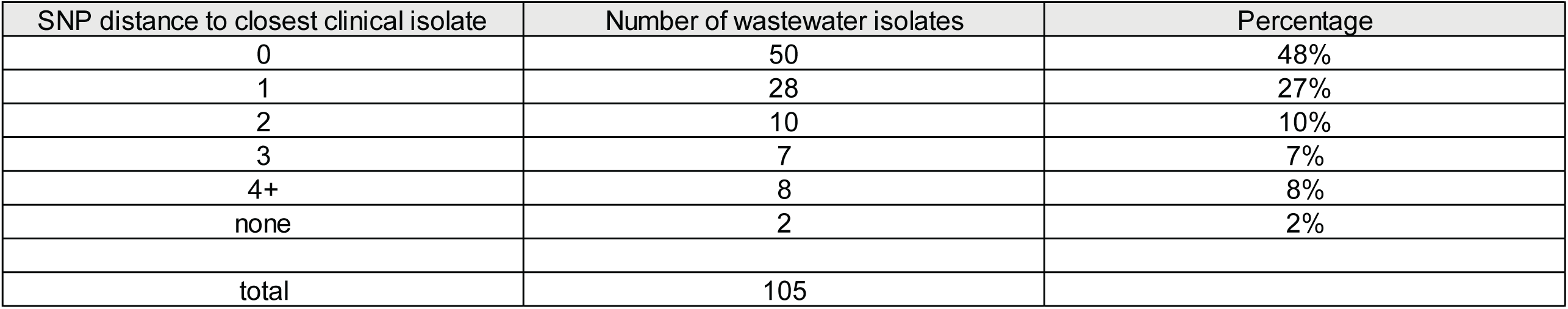
SNP distance to closest clinical isolates.

| SNP distance to closest clinical isolate | Number of wastewater isolates | Percentage |
| --- | --- | --- |
| 0 | 50 | 48% |
| 1 | 28 | 27% |
| 2 | 10 | 10% |
| 3 | 7 | 7% |
| 4+ | 8 | 8% |
| none | 2 | 2% |
| total | 105 |  |

## Notes

### Competing Interest Statement

The authors have declared no competing interest.

## References

1 Satoh, K. et al. Candida auris sp. nov., a novel ascomycetous yeast isolated from the external ear canal of an inpatient in a Japanese hospital. Microbiol Immunol 53, 41–44 (2009). 10.1111/j.1348-0421.2008.00083.x

2 O’Meara, T. R. Understanding pathogen emergence through the lens of Candida auris. Nat Microbiol (2024). 10.1038/s41564-024-01700-2

3 McCormick, T. S. & Ghannoum, M. Time to Think Antifungal Resistance Increased Antifungal Resistance Exacerbates the Burden of Fungal Infections Including Resistant Dermatomycoses. Pathog Immun 8, 158–176 (2023). 10.20411/pai.v8i2.656

4 Chowdhary, A., Jain, K. & Chauhan, N. Candida auris Genetics and Emergence. Annu Rev Microbiol 77, 583–602 (2023). 10.1146/annurev-micro-032521-015858

5 Eix, E. F. & Nett, J. E. Candida auris: Epidemiology and Antifungal Strategy. Annu Rev Med (2024). 10.1146/annurev-med-061523-021233

6 Chow, N. A. et al. Tracing the Evolutionary History and Global Expansion of Candida auris Using Population Genomic Analyses. mBio 11 (2020). 10.1128/mBio.03364-19

7 Burrack, L. S., Todd, R. T., Soisangwan, N., Wiederhold, N. P. & Selmecki, A. Genomic Diversity across Candida auris Clinical Isolates Shapes Rapid Development of Antifungal Resistance In Vitro and In Vivo. mBio 13, e0084222 (2022). 10.1128/mbio.00842-22

8 CDC. Antibiotic Resistance Threats in the United States, 2019, <https://www.cdc.gov/antimicrobial-resistance/data-research/threats/> (2024).

9 Egger, N. B. et al. The rise of Candida auris: from unique traits to co-infection potential. Microb Cell 9, 141–144 (2022). 10.15698/mic2022.08.782

10 Benedict, K., Forsberg, K., Gold, J. A. W., Baggs, J. & Lyman, M. Candida auris‒ Associated Hospitalizations, United States, 2017-2022. Emerg Infect Dis 29, 1485–1487 (2023). 10.3201/eid2907.230540

11 Soriano, A. et al. Invasive candidiasis: current clinical challenges and unmet needs in adult populations. J Antimicrob Chemother 78, 1569–1585 (2023). 10.1093/jac/dkad139

12 Pappas, P. G., Lionakis, M. S., Arendrup, M. C., Ostrosky-Zeichner, L. & Kullberg, B. J. Invasive candidiasis. Nat Rev Dis Primers 4, 18026 (2018). 10.1038/nrdp.2018.26

13 Cortegiani, A. et al. Epidemiology, clinical characteristics, resistance, and treatment of infections by Candida auris. J Intensive Care 6, 69 (2018). 10.1186/s40560-018-0342-4

14 Suphavilai, C. et al. Detection and characterisation of a sixth Candida auris clade in Singapore: a genomic and phenotypic study. Lancet Microbe 5, 100878 (2024). 10.1016/S2666-5247(24)00101-0

15 Salam, M. A. et al. Antimicrobial Resistance: A Growing Serious Threat for Global Public Health. Healthcare (Basel) 11 (2023). 10.3390/healthcare11131946

16 Barber, C. et al. Community-Scale Wastewater Surveillance of Candida auris during an Ongoing Outbreak in Southern Nevada. Environ Sci Technol 57, 1755–1763 (2023). 10.1021/acs.est.2c07763

17 Rossi, A. et al. Candida auris Discovery through Community Wastewater Surveillance during Healthcare Outbreak, Nevada, USA, 2022. Emerg Infect Dis 29, 422–425 (2023). 10.3201/eid2902.221523

18 Babler, K. et al. Detection of the clinically persistent, pathogenic yeast spp. Candida auris from hospital and municipal wastewater in Miami-Dade County, Florida. Sci Total Environ 898, 165459 (2023). 10.1016/j.scitotenv.2023.165459

19 Zulli, A. et al. Prospective study of Candida auris nucleic acids in wastewater solids in 190 wastewater treatment plants in the United States suggests widespread occurrence. mBio 15, e0090824 (2024). 10.1128/mbio.00908-24

20 Chavez, J. et al. Early Introductions of Candida auris Detected by Wastewater Surveillance, Utah, USA, 2022-2023. Emerg Infect Dis 30, 2107–2117 (2024). 10.3201/eid3010.240173

21 Harrington, A. et al. Urban monitoring of antimicrobial resistance during a COVID-19 surge through wastewater surveillance. Sci Total Environ 853, 158577 (2022). 10.1016/j.scitotenv.2022.158577

22 Vo, V. et al. Identification and genome sequencing of an influenza H3N2 variant in wastewater from elementary schools during a surge of influenza A cases in Las Vegas, Nevada. Sci Total Environ 872, 162058 (2023). 10.1016/j.scitotenv.2023.162058

23 Vo, V. et al. Detection of the Omicron BA.1 Variant of SARS-CoV-2 in Wastewater From a Las Vegas Tourist Area. JAMA Netw Open 6, e230550 (2023). 10.1001/jamanetworkopen.2023.0550

24 Ambrosio, F. J., 3rd et al. TheiaEuk: a species-agnostic bioinformatics workflow for fungal genomic characterization. Front Public Health 11, 1198213 (2023). 10.3389/fpubh.2023.1198213

25 Gorzalski, A. et al. The use of whole-genome sequencing and development of bioinformatics to monitor overlapping outbreaks of Candida auris in southern Nevada. Front Public Health 11, 1198189 (2023). 10.3389/fpubh.2023.1198189

26 Haas, G. Infections from superbug C. auris declining in Southern Nevada, state data shows, <www.8newsnow.com/news/local-news/infections-from-superbug-c-auris-declining-in-southern-nevada-state-data-shows/> (2024).

27 Hynes, M. ‘Superbug’ fungus cases hit record high in Southern Nevada, <https://www.reviewjournal.com/life/health/superbug-fungus-cases-hit-record-high-in-southern-nevada-3005044/> (2024).

28 Nguyen, H. et al. CCPA: Cloud-based, self-learning modules for Consensus Pathway Analysis using GO, KEGG and Reactome. Briefings in Bioinformatics (2024). 10.1093/bib/bbae222

29 Nguyen, H. et al. CPA: A web-based platform for Consensus Pathway Analysis and interactive visualization. Nucleic Acids Research 49, W114--W124 (2021).

30 Harrington, A. et al. Environmental Surveillance of Flood Control Infrastructure Impacted by Unsheltered Individuals Leads to the Detection of SARS-CoV-2 and Novel Mutations in the Spike Gene. Environ Sci Technol Lett 11, 410–417 (2024). 10.1021/acs.estlett.3c00938

31 Barlam, T. F. et al. Implementing an Antibiotic Stewardship Program: Guidelines by the Infectious Diseases Society of America and the Society for Healthcare Epidemiology of America. Clin Infect Dis 62, e51–77 (2016). 10.1093/cid/ciw118

32 Arastehfar, A. et al. Echinocandin persistence directly impacts the evolution of resistance and survival of the pathogenic fungus Candida glabrata. mBio 15, e0007224 (2024). 10.1128/mbio.00072-24

33 Larsson, D. G. J. & Flach, C. F. Antibiotic resistance in the environment. Nat Rev Microbiol 20, 257–269 (2022). 10.1038/s41579-021-00649-x

34 Huang, F. et al. Peroxisome disruption alters lipid metabolism and potentiates antitumor response with MAPK-targeted therapy in melanoma. J Clin Invest 133 (2023). 10.1172/JCI166644

35 Zhuang, X. et al. Early Detection of Novel SARS-CoV-2 Variants from Urban and Rural Wastewater through Genome Sequencing and Machine Learning. medRxiv (2024). 10.1101/2024.04.18.24306052

36 Zhuang, X. et al. Drug Use Patterns in Wastewater and Socioeconomic and Demographic Indicators. JAMA Netw Open 7, e2432682 (2024). 10.1001/jamanetworkopen.2024.32682

37 Vo, V. et al. Use of wastewater surveillance for early detection of Alpha and Epsilon SARS-CoV-2 variants of concern and estimation of overall COVID-19 infection burden. Sci Total Environ 835, 155410 (2022). 10.1016/j.scitotenv.2022.155410

38 Quick, J. et al. Multiplex PCR method for MinION and Illumina sequencing of Zika and other virus genomes directly from clinical samples. Nat Protoc 12, 1261–1276 (2017). 10.1038/nprot.2017.066

39 Wang, M. X., et al. Olivar: towards automated variant aware primer design for multiplex tiled amplicon sequencing of pathogens. Nat Commun 15, 6306 (2024). 10.1038/s41467-024-49957-9

40 Xie, N. G. et al. Designing highly multiplex PCR primer sets with Simulated Annealing Design using Dimer Likelihood Estimation (SADDLE). Nat Commun 13, 1881 (2022). 10.1038/s41467-022-29500-4

41 Chen, S., Zhou, Y., Chen, Y. & Gu, J. fastp: an ultra-fast all-in-one FASTQ preprocessor. Bioinformatics 34, i884–i890 (2018). 10.1093/bioinformatics/bty560

42 Li, H. & Durbin, R. Fast and accurate long-read alignment with Burrows-Wheeler transform. Bioinformatics 26, 589–595 (2010). 10.1093/bioinformatics/btp698

43 Li, H. et al. The Sequence Alignment/Map format and SAMtools. Bioinformatics 25, 2078–2079 (2009). 10.1093/bioinformatics/btp352

44 Grubaugh, N. D. et al. An amplicon-based sequencing framework for accurately measuring intrahost virus diversity using PrimalSeq and iVar. Genome Biol 20, 8 (2019). 10.1186/s13059-018-1618-7

45 Cingolani, P. et al. A program for annotating and predicting the effects of single nucleotide polymorphisms, SnpEff: SNPs in the genome of Drosophila melanogaster strain w1118; iso-2; iso-3. Fly (Austin) 6, 80–92 (2012). 10.4161/fly.19695

46 Bagal, U. R. et al. MycoSNP: A Portable Workflow for Performing Whole-Genome Sequencing Analysis of Candida auris. Methods Mol Biol 2517, 215–228 (2022). 10.1007/978-1-0716-2417-3_17

47 Ewels, P. A. et al. The nf-core framework for community-curated bioinformatics pipelines. Nat Biotechnol 38, 276–278 (2020). 10.1038/s41587-020-0439-x

48 Quinlan, A. R. & Hall, I. M. BEDTools: a flexible suite of utilities for comparing genomic features. Bioinformatics 26, 841–842 (2010). 10.1093/bioinformatics/btq033

49 Marcais, G. et al. MUMmer4: A fast and versatile genome alignment system. PLoS Comput Biol 14, e1005944 (2018). 10.1371/journal.pcbi.1005944

50 Lo, C. C. & Chain, P. S. Rapid evaluation and quality control of next generation sequencing data with FaQCs. BMC Bioinformatics 15, 366 (2014). 10.1186/s12859-014-0366-2

51 McKenna, A. et al. The Genome Analysis Toolkit: a MapReduce framework for analyzing next-generation DNA sequencing data. Genome Res 20, 1297–1303 (2010). 10.1101/gr.107524.110

52 Cingolani, P. et al. Using Drosophila melanogaster as a Model for Genotoxic Chemical Mutational Studies with a New Program, SnpSift. Front Genet 3, 35 (2012). 10.3389/fgene.2012.00035

53 broadinstitute/broad-fungalgroup. <github.com/broadinstitute/broad-fungalgroup/tree/master/scripts/SNPs> (

54 Kozlov, A. M., Darriba, D., Flouri, T., Morel, B. & Stamatakis, A. RAxML-NG: a fast, scalable and user-friendly tool for maximum likelihood phylogenetic inference. Bioinformatics 35, 4453–4455 (2019). 10.1093/bioinformatics/btz305

55 Skrzypek, M. S. et al. The Candida Genome Database (CGD): incorporation of Assembly 22, systematic identifiers and visualization of high throughput sequencing data. Nucleic Acids Res 45, D592–D596 (2017). 10.1093/nar/gkw924

56 Patro, R., Duggal, G., Love, M. I., Irizarry, R. A. & Kingsford, C. Salmon provides fast and bias-aware quantification of transcript expression. Nat Methods 14, 417–419 (2017). 10.1038/nmeth.4197

57 Pertea, M. et al. StringTie enables improved reconstruction of a transcriptome from RNA-seq reads. Nat Biotechnol 33, 290–295 (2015). 10.1038/nbt.3122

58 Kanehisa, M., Furumichi, M., Tanabe, M., Sato, Y. & Morishima, K. KEGG: new perspectives on genomes, pathways, diseases and drugs. Nucleic Acids Research 45, D353–D361 (2017).

59 The Gene Ontology Consortium. The Gene Ontology resource: enriching a GOld mine. Nucleic Acids Research 49, D325–D334 (2021).

60 Blast, G. PSI-BLAST: a new generation of protein database search programs. Nucleic Acids Res 25, 3389–3402 (1997).

61 Camacho, C. et al. BLAST+: architecture and applications. BMC bioinformatics 10, 1–9 (2009).

62 Nguyen, H. et al. RCPA: An Open-Source R Package for Data Processing, Differential Analysis, Consensus Pathway Analysis, and Visualization. Current Protocols 4, e1036 (2024).

63 Korotkevich, G. et al. Fast gene set enrichment analysis. BioRxiv, 060012 (2021).

64 Benjamini, Y. & Hochberg, Y. Controlling the false discovery rate: a practical and powerful approach to multiple testing. Journal of the Royal Statistical Society: Series B (Methodological) 57, 289--300 (1995).

